# A pretreatment T cell signalling score identifies clinical pembrolizumab response in non-small cell lung cancer patients

**DOI:** 10.64898/2026.02.04.26345546

**Authors:** Judith D.J. Verdonk, Rob ter Heine, Berber Piet, Esther van Rijssen, Michel M. van den Heuvel, Hans J.P.M. Koenen, Ruben L. Smeets, the DEDICATION consortium

## Abstract

**Background:** Immune checkpoint inhibitors (ICIs) targeting the programmed death (ligand)-1 (PD-1/PD-L1) axis, like pembrolizumab, have significantly improved survival in non-small cell lung cancer (NSCLC). However, less than 50% of patients respond. Identifying early-response biomarkers is crucial to personalize therapy, thereby preventing ineffective, expensive and potentially harmful treatment.

**Methods:** We applied a novel *ex vivo* immunopharmacological bioassay to assess pembrolizumab-dependent T cell signalling in baseline peripheral blood mononuclear cells (PBMCs) from 64 NSCLC patients. PBMCs were stimulated with anti-CD3/CD28 with or without pembrolizumab, and phosphorylation states of PD-1-dependent T cell receptor (TCR) signalling pathways were measured by spectral flow cytometry. A composite signalling score was calculated representing the net pembrolizumab-induced phosphorylation response and patients were classified as low, optimal and high modulation responders based on this signalling score. Associations with progression-free survival and overall survival (OS) were evaluated using univariate Cox regression.

**Results:** Patients with optimal baseline pembrolizumab-induced signalling scores exhibited significantly higher signalling score outcomes than those with low modulation (*p* < 0.0001) and lower than patients with excessive modulation (*p* < 0.01) and had significantly longer OS (HR = 2.83, *p* = 0.013; and HR = 12.05, *p* = 0.003, respectively). Notably, conventional pharmacodynamic parameters, including half-maximal effective concentration (EC_50_) for PD-1 receptor occupancy and maximum IL-2 production (E_max_), were not associated with clinical outcomes, underscoring the unique predictive value of the phosphorylation-based signalling score. *In vivo*, pembrolizumab-induced T cell activation changes and TCR signalling inhibition post-treatment correlated with shorter survival (HRs = 1.33-1.95), consistent with our *ex vivo* findings.

**Conclusions:** We demonstrate that a pretreatment signalling score derived from *ex vivo* pembrolizumab-modulated T cell phosphorylation identifies clinical response in NSCLC. This functional bioassay offers a novel approach to identify patients most likely to benefit from ICI therapy, potentially enabling personalised treatment decisions before therapy initiation.

**Graphical abstract text:** Our findings reveal that pretreatment, pembrolizumab-dependent modulation of T cell phosphorylation identifies clinical response in NSCLC. Furthermore, we introduce an overall signalling score, reflecting the net phosphorylation profile, which could serve as a potential predictive biomarker to distinguish responders from non-responders, thereby supporting biomarker-driven therapeutic strategies.

## 1. Background

Immune checkpoint inhibitors (ICIs) have greatly improved survival of cancer, including non-small cell lung cancer (NSCLC). [1, 2] By targeting the programmed death (ligand)-1 (PD-1/PD-L1) axis with specific ICIs, T cell-mediated immunity can be restored, and cancer immune evasion can be overcome.

PD-1 inhibitor pembrolizumab is currently first-line treatment for advanced-stage NSCLC. However, less than half of patients respond to anti-PD-1 therapy. [3] Consequently, more than half of patients receiving immunotherapy will not have a clinically relevant benefit but are nevertheless exposed to potential immunotherapy side-effects and might lose the opportunity to benefit from effective treatment alternatives. On top of that, the exponentially increasing costs of new drugs, like ICIs, endanger population-wide accessibility to life-saving treatments.

Despite the urgent need, reliable biomarkers to identify patients who will benefit from anti-PD-1 therapy remain lacking. In NSCLC, PD-L1 expression on tumour tissue assessed by immunohistochemistry and radiological evaluation via computed tomography (CT) are the standard approaches for predicting and monitoring treatment response. [3] Yet, both methods offer only modest predictive accuracy, leaving precision immunotherapy in its infancy. [4-6] Therefore, the development of robust predictive biomarkers for ICI response has become a major focus in cancer research worldwide.

Measurement of peripheral blood immune cell composition provides a minimally invasive biomarker for clinical application. Various parameters, including white blood cell ratios, circulating immune cell subsets and their functional states, cytokines, soluble checkpoint molecules, autoantibodies, and emerging tumour-derived circulating biomarkers, have shown associations with immunotherapy response. Nevertheless, their individual predictive accuracy remains insufficient to reliably predict treatment response. [7, 8] Identifying robust early-response biomarkers is therefore essential to enable personalised therapy, thereby preventing ineffective, expensive and potentially harmful treatment.

To address this gap, we aimed to comprehensively characterise the functional phenotype of T cells in NSCLC patients receiving pembrolizumab. We hypothesised that the functional and therapy-modulated T cell profiles may predict response to ICI therapy. Previously, we developed an *ex vivo* immuno-pharmacological bioassay to assess PD-1 receptor pharmacology. [9] This bioassay evaluates ICI pharmacodynamics of at four levels: target binding, intracellular T cell signalling, T cell phenotype and effector function (Fig. 1). Here, we sought to identify covariates across these levels that associate with clinical pembrolizumab response, aiming to define a functional T cell signature predictive of ICI efficacy.

**Fig. 1.**
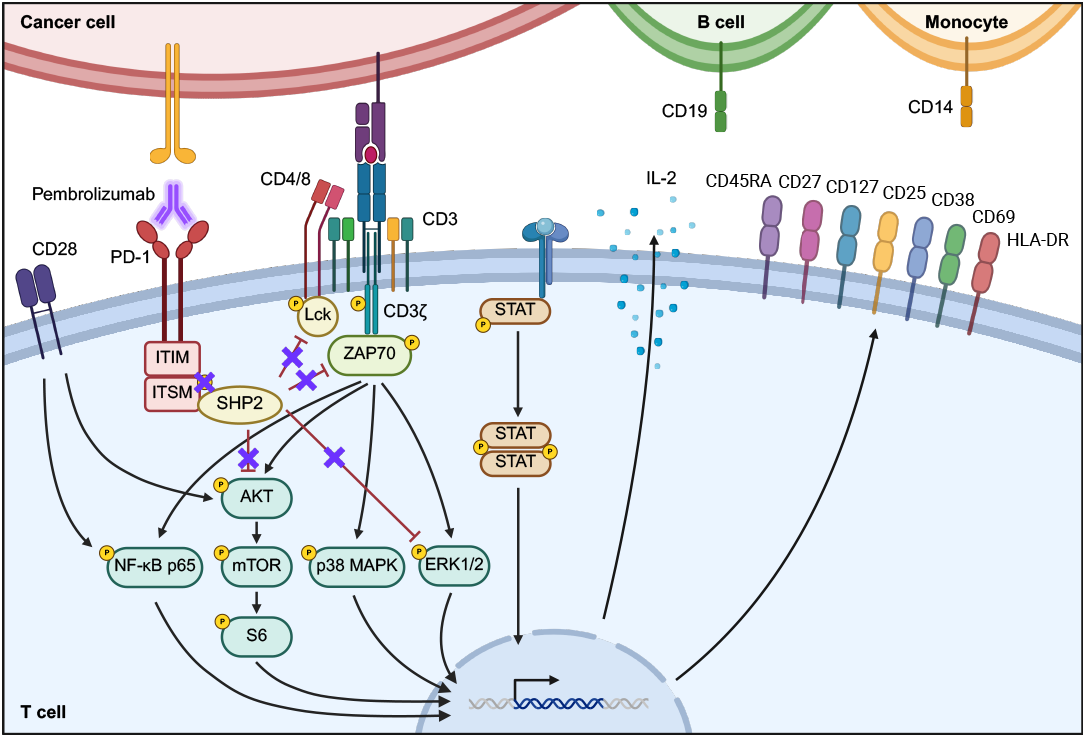
Functional T cell phenotype evaluation using an *ex vivo* immunopharmacological bioassay for immuno-pharmacodynamics of immune checkpoint inhibitors. The functional T cell phenotype was evaluated at four levels: (1) target binding, (2) intracellular T cell signalling, (3) T cell phenotype, and (4) effector function. For target binding, expression of PD-1 was measured. For intracellular T cell signalling, phosphorylation of proximal signalling molecules PD-1(Y248), Lck(Y394), Lck(Y05), CD3ζ(Y142), ZAP70(Y319)/Syk(Y352), and ZAP70(Y292) were assessed, as well as phosphorylation of downstream effectors NF-κB p65(S529), AKT(T308), mTOR(S2448), S6(S235/S236), p38 MAPK(T180/Y182), ERK1/2(T202/Y204), STAT1(Y701), STAT3(Y705), STAT5(Y694), and STAT6(Y641). The phenotypical level was characterised by, lineage markers CD3, CD4, CD8, CD14, CD19, CD25, CD27, CD45RA, and CD127, alongside maturation markers CD27 and CD45RA, as well as activation markers CD25, CD38, CD69, and HLA-DR, and senescence markers CD28 and PD-1. Lastly, IL-2 was measured as a surrogate marker for T cell effector function. PD-1 = programmed cell death protein 1, Lck = leukocyte-specific tyrosine kinase, CD = cluster of differentiation, ZAP70 = zeta-chain-associated protein kinase-70, Syk = spleen tyrosine kinase, NF-κB = nuclear factor-κB p65 subunit, mTOR = mammalian target of rapamycin, MAPK = mitogen-activated protein kinase, ERK = extracellular signal-regulated kinase, STAT = signal transducer and activator of transcription, HLA-DR = human leukocyte antigen – DR isotype, IL-2 = interleukin-2.

## 2. Methods

### 2.1. Study population

A total of 64 patients with advanced-stage NSCLC were enrolled in this study. All patients participated in the prospective DEDICATION-1 trial (ClinicalTrials.gov ID: NCT04909684) and were recruited in the Radboudumc in Nijmegen, The Netherlands. This study was conducted in accordance with the Medical Research Involving Human Subjects Act (WMO) and received approval from the Medical Ethics Committee Oost-Nederland (METC Oost-Nederland, Nijmegen, The Netherlands). Written informed consent was obtained from all participants prior to inclusion.

### 2.2. Response evaluation

Therapy response was assessed in terms of progression free survival (PFS) and overall survival (OS). PFS was defined as the time from the start of pembrolizumab therapy to disease progression, evaluated according to the standard-of-care schedule at baseline (BL), six weeks (6w), twelve weeks and every three months thereafter. Patients were censored for PFS if they were alive without disease progression at the data cut-off (February 2025), were lost to follow-up or had died from unknown causes without documented evidence of disease progression. OS was specified as the time from the start of pembrolizumab therapy to death from any cause. Patients were censored for OS if they were lost to follow-up or alive at data cut-off. Follow-up duration was defined as the time from initiation of pembrolizumab therapy to loss to follow-up, death or data cut-off.

### 2.3. Sample collection

Peripheral blood samples were obtained at BL and at three weeks (3w) and 6w after start of pembrolizumab treatment. As part of routine care platelet, lymphocyte, neutrophil and monocyte count were determined from whole blood to obtain platelet to lymphocyte ratio, neutrophil to lymphocyte ratio, and monocyte to lymphocyte ratio. For research purposes whole blood was collected in 10 mL EDTA tubes (BD, NJ, USA) and processed within four hours. Peripheral blood mononuclear cells (PBMCs) were isolated from 7 mL whole blood using the AutoMACS whole blood PBMC isolation kit (Miltenyi Biotech, Bergisch Gladbach, Germany) according to manufacturer’s instructions. Cells were cryopreserved in 1 mL freezing medium (Gibco® RPMI 1640 Dutch modified, Thermo Fisher Scientific, Waltham, USA; addition of 1 % sodium pyruvate, 1 % glutamax, 1 % penicillin and streptomycin and 10 % dimethyl sulfoxide, all Thermo Fisher Scientific; 10 % human pooled serum, manufactured in-house) in liquid nitrogen until further use.

### 2.4. *Ex vivo* immunopharmacological bioassay

After thawing, the PBMCs were used in an *ex vivo* immunopharmacological bioassay, as described previously. [9] In brief, the functional phenotype of T cells was measured either directly after thawing or following *in vitro* stimulation of the PBMCs. Prior to *in vitro* stimulation, PBMCs were first incubated for 30 min at 37 °C, 5 % CO2, 95 % humidity with a concentration range of pembrolizumab (0, 0.000025, 0.00025, 0.0025, 0.025, 2.5, 25 or 250 μg/mL; Keytruda® 25 mg/mL, Merck & Co, NJ, USA) in fresh culture medium (Gibco® RPMI 1640 Dutch modified; supplemented with 5 % Foetal bovine serum (FBS), Greiner Bio-One; 1 mM pyruvate, 2 mM glutamax, 100 U/mL penicillin, and 100 mg/mL streptomycin, all Thermo Fisher Scientific), mimicking *in vivo* pembrolizumab concentrations. [10, 11] Subsequently, cells were stimulated with 0.1 μg/mL soluble aCD3 (Purified NA/LE Mouse Anti-Human CD3, OKT3, BD Biosciences, Franklin Lakes, NJ, USA) and 1.0 μg/mL soluble aCD28 (Purified NA/LE Mouse Anti-Human CD28, CD28.2, BD Biosciences) for 48 hours at 37 °C, 5 % CO2, 95 % humidity, while pembrolizumab was still present. The functional T cell phenotype was then analysed at four levels, as described below (2.4.1-2.4.4).

#### 2.4.1. Phenotypical assessment using spectral flow cytometry

In short, the phenotype of PBMCs was assessed using spectral flow cytometry, using the flow cytometry panel for surface staining listed in Table S1. This panel was adapted from the OMIP-112 panel described by Waaijer *et al*. [12] to fit the specific requirements of this study. Pembrolizumab, bound to the PD-1 receptor on the cell surface was measured using a secondary IgG4 Fc – PE antibody (SouthernBiotech, Alabama, USA), which binds the Fc tail of pembrolizumab. Samples were measured on a spectral flow cytometer (SONY ID7000, Minato-Ku, Tokyo, Japan), and data were analysed using Kaluza Analysis Software (version 2.2.0, Beckman Coulter, Indianapolis, Indiana, USA). The gating strategy for the immune cell subtypes is illustrated in Fig. S1.

#### 2.4.2. Receptor occupancy measurement using spectral flow cytometry

From the obtained phenotype data, PD-1 expression on PD-1 positive T cells was used to calculate receptor occupancy. The following formula, obtained from Shchelokov and Demin [13], which uses free PD-1 receptors (R_Free_) and PD-1 receptors measured pre-dose (R_Pre-dose_), was used:

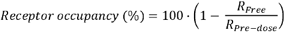

Background mean fluorescence intensity (MFI) for PD-1 expression was measured using samples with full PD-1 blockage by pembrolizumab and subtracted from MFI of PD-1 receptors measured in all samples to obtain R_Free_. MFI of R_Pre-dose_ was obtained from samples stimulated in the absence of pembrolizumab after correction for background MFI.

#### 2.4.3. Assessment of phosphorylation levels of T cell signalling markers using spectral flow cytometry

As previously described [9], to assess phosphorylation levels of T cell signalling proteins, cells were intracellularly stained using a panel for phospho-staining, targeting proximal kinases (leukocyte-specific tyrosine kinase (Lck), and zeta-chain-associated protein kinase-70 (ZAP70)), as well as downstream effectors (nuclear factor-κB p65 subunit (NF-κB p65), AKT, mammalian target of rapamycin (mTOR), S6, p38 mitogen-activated protein kinase (MAPK), extracellular signal-regulated kinase (ERK) 1/2, and signal transducer and activator of transcription (STAT) proteins). The anti-PD-1(Y248) antibody, used for measurement of the phosphorylation level of the intracellular tail of PD-1, was a kind gift of Beth Israel Deaconess Medical Center, Harvard Medical School, Boston. [14] The complete panel is listed in Table S1. Samples were analysed using spectral flow cytometry, as described above.

#### 2.4.4. Assessment of IL-2 production potential using Luminex

Briefly, interleukin-2 (IL-2) concentration in culture supernatant was determined using MILLIPLEX® Human Cytokine/ Chemokine/Growth Factor Panel A kit (Merck Millipore, Darmstadt, Germany) according to manufacturer’s instructions and measured using a FLEXMAP 3D system (Luminex Corporation). Data were analysed using Bio-Plex Manager software (version 6.2.0.175, Bio-Rad Laboratories, Hercules, California, USA).

### 2.5. Modelling of PD-1 receptor occupancy and IL-2 dose-response curves

The obtained data describing receptor occupancy and absolute IL-2 concentrations were analysed by means of non-linear mixed effects modelling using the software package NONMEM 7.51 with the first order conditional estimation method with interaction (FOCE-I). The pembrolizumab concentrations were used as independent variable and the receptor occupancy and absolute IL-2 concentrations as dependent variables in two independent analyses. Parameter uncertainty (relative standard error of estimates - RSE) was assessed using the covariance step in NONMEM.

The underlying assumptions for the non-linear mixed effects modelling analysis were that 1) the relationship could be described with a sigmoidal maximum effect (E_max_) model and that 2) inter-individual variability was log-normally distributed. Residual error was described by means of additive and/or proportional error models, depending on the data. Model building was guided by standard goodness-of-fit plots and model identifiability, as assessed with model stability as well as the condition number and parameter correlation obtained from the covariance step.

The following equations were fitted to the data:

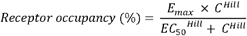

In this equation, E_max_ is the maximum receptor occupancy, which was assumed (and fixed) to be 100%, C is the pembrolizumab concentration in µg/mL, EC_50_ is the concentration at which 50% receptor occupancy is expected and Hill is the hill coefficient, describing the steepness of the exposure response relationship.

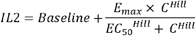

In this equation, Baseline is the baseline IL-2 concentration (pg/mL), E_max_ is the maximum IL-2 concentration (pg/mL), C is the pembrolizumab concentration in µg/mL, EC_50_ is the concentration at which 50% of the effect is expected and Hill is the Hill coefficient, describing the steepness of the exposure response relationship.

The obtained parameter estimates were used to simulate receptor occupancy and IL-2 concentrations for 1,000 individuals. The 10^th^, 50^th^ and 90^th^ percentiles of simulated values were used for data visualisation.

### 2.6. Statistical analysis

Statistical analyses were performed using R (version 2024.04.02). Median PFS and OS were obtained from Kaplan Meier curves at 50% survival probability. Associations of covariates with PFS and OS and corresponding hazard ratios (HRs) were examined using univariate Cox regression on z-scaled continuous covariate data. For continuous variables, the linearity assumption was evaluated both statistically, through natural spline models, and graphically, by plotting the estimated log-HRs against the covariates. Although some minor non-linear trends were observed, no adjustments were made in the final models, as the present analysis is exploratory in nature. To explore patterns in immune profiles, heatmaps were generated using Euclidean distance and Ward.D2 hierarchical clustering on z-scaled data. HRs for patient groups were calculated using univariate Cox regression, with the group exhibiting the longest median survival time used as the reference group. Correlation network analysis was conducted using Spearman’s rank correlation coefficient (*ρ*) on z-scaled data, with correlations considered biologically relevant if *ρ* > 0.7 and *p* < 0.05. To summarise pembrolizumab-dependent signalling patterns per patient, an overall signalling score was calculated using weighted gene co-expression network analysis. Eigengenes were extracted for both the signalling markers expected to be involved in a tumour supporting effect (Lck(Y505), ZAP70(Y292), MAPK(Thr180/Tyr182), and STAT6(Y641)), as well as for the other signalling markers, which are expected to be involved in an anti-tumour effect. [14-22] The signalling score was defined as the difference between these eigengenes, representing the net pembrolizumab-dependent signalling profile. Group comparisons were performed using the Wilcoxon rank-sum test, due to the small sample size in one modulation response group (n = 2). For receptor occupancy and IL-2 analysis, the full pembrolizumab concentration range was used. For further analysis of T cell phenotype and signalling following *in vitro* stimulation, PBMCs were either left unstimulated (Unstim), stimulated in the absence of pembrolizumab (Stim) or stimulated in the presence of 2.5 or 25 µg/mL pembrolizumab (Pembro). Changes in phosphorylation levels of T cell signalling markers were quantified by subtracting MFI in Unstim samples from MFI in Stim samples (ΔStim) and by subtracting MFI in Stim samples from MFI in Pembro samples (ΔPembro).

## 3. Results

### 3.1. Patient and treatment characteristics

Blood samples from three timepoints of 64 advanced-stage NSCLC patients were analysed. Patient and treatment characteristics are shown in Table 1. In brief, 39.1% of patients were female and 60.9% were male. Median age at study enrolment was 67 years (range 31 – 83). The majority (62.5%) of patients had PD-L1 expression < 50%. Almost all patients had a smoking history, 20.3% were current and 75.0% former smokers. Patients received either pembrolizumab monotherapy (26.6%), combination therapy of pembrolizumab, pemetrexed and platinum (Pembro-pem-plat; 53.1%), or combination therapy of pembrolizumab, paclitaxel and platinum (Pembro-pacli-plat; 20.3%). All patients were immunotherapy naive and most patients received no other lung cancer treatment in the three months prior to study enrolment (95.3%). The other 4.7% underwent surgery within three months before study enrolment.

**Table 1.**
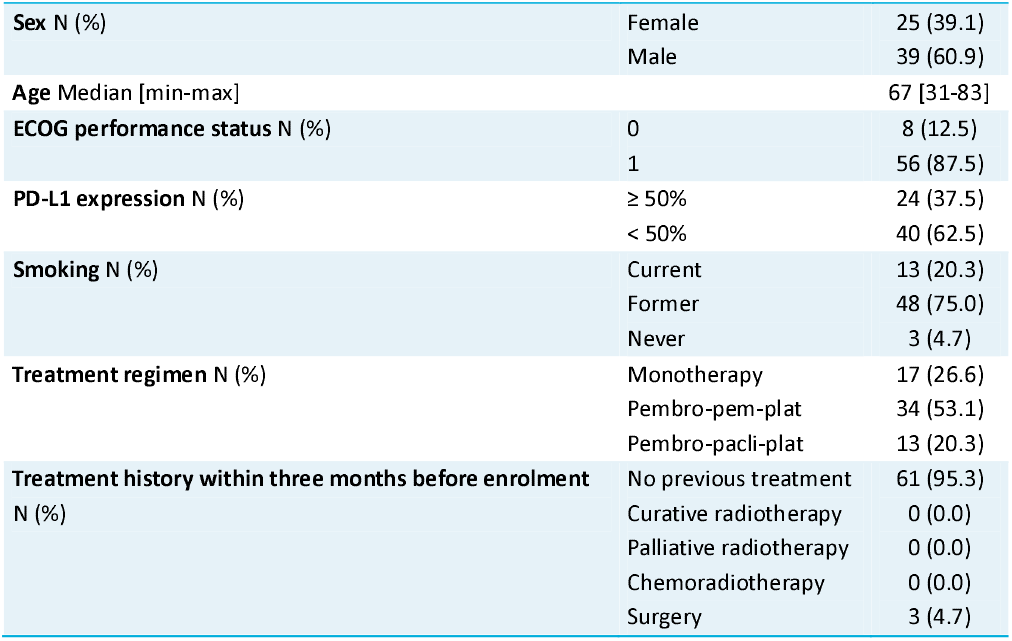
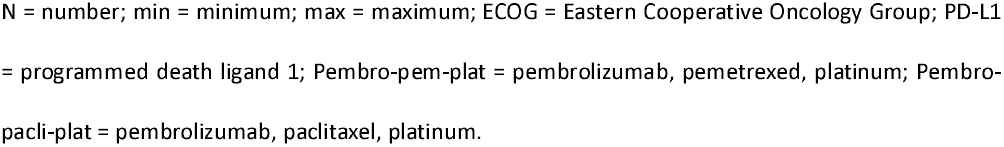
Patient and treatment characteristics.

|  |  |  |
| --- | --- | --- |
| **Sex** N (%) | Female | 25 (39.1) |
|  | Male | 39 (60.9) |
| **Age** Median [min-max] |  | 67 [31-83] |
| **ECOG performance status** N (%) | 0 | 8 (12.5) |
|  | 1 | 56 (87.5) |
| **PD-L1 expression** N (%) | ≥ 50% | 24 (37.5) |
|  | < 50% | 40 (62.5) |
| **Smoking** N (%) | Current | 13 (20.3) |
|  | Former | 48 (75.0) |
|  | Never | 3 (4.7) |
| **Treatment regimen** N (%) | Monotherapy | 17 (26.6) |
|  | Pembro-pem-plat | 34 (53.1) |
|  | Pembro-pacli-plat | 13 (20.3) |
| **Treatment history within three months before enrolment** N (%) | No previous treatment | 61 (95.3) |
|  | Curative radiotherapy | 0 (0.0) |
|  | Palliative radiotherapy | 0 (0.0) |
|  | Chemoradiotherapy | 0 (0.0) |
|  | Surgery | 3 (4.7) |

N = number; min = minimum; max = maximum; ECOG = Eastern Cooperative Oncology Group; PD-L1 = programmed death ligand 1; Pembro-pem-plat = pembrolizumab, pemetrexed, platinum; Pembro-pacl-plat = pembrolizumab, paclitaxel, platinum.

### 3.2. Survival

Kaplan Meier survival curves of the total study population for PFS and OS are shown in Fig. 2A-B. Median PFS and OS were 248 and 538 days respectively.

**Fig. 2.**
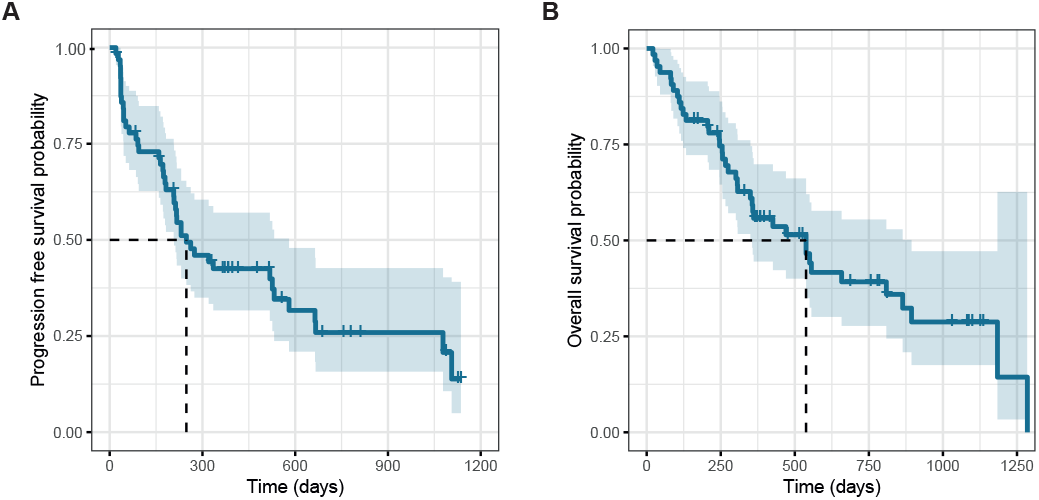
Kaplan Meier survival curves showing (A) PFS and (B) OS in the total study population. The survival curves of 64 NSCLC patients are shown with 95% confidence interval. Median PFS and OS were 248 days and 538 days respectively. PFS = progression free survival, OS = overall survival, NSCLC = non-small cell lung cancer.

### 3.3. Functional T cell profile

To identify a functional T cell profile which is predictive of anti-PD-1 treatment response, the effect of pembrolizumab on T cells was studied at four levels: 1) PD-1 receptor occupancy; 2) T cell signalling; 3) T cell phenotype and 4) IL-2 production potential.

#### 3.3.1. PD-1 receptor occupancy and IL-2 induction potential are not associated with PFS and OS

First, the effect of increasing pembrolizumab concentrations on PD-1 receptor occupancy and IL-2 induction was studied. For both PD-1 receptor occupancy and IL-2 induction in response to increasing pembrolizumab concentrations, *in vitro* measured data (n = 60 for receptor occupancy and n = 61 for IL-2 concentration) with corresponding *in silico* modelled concentration-response curves are shown in Fig. 3A-B. Corresponding parameter estimates are listed in Table S2 and the goodness-of-fit plots and typical curves are shown in Fig. S2A-D. Both PD-1 receptor occupancy and IL-2 concentration increased pembrolizumab concentration-dependently. The half maximal effective concentration (EC_50_) of pembrolizumab for PD-1 receptor occupancy was 0.0018 µg/mL, whereas the EC_50_ of pembrolizumab for induction of IL-2 concentration was 12.14 µg/mL (over 6500-fold higher than the EC_50_ for receptor occupancy). Patient variation for EC_50_ of PD-1 receptor occupancy (range 0.00014 – 0.0065 µg/mL) and E_max_ for IL-2 concentration (range 27.56 – 1737.70 pg/mL) was observed, as shown in Fig. 3C-D. Hazard ratio analyses indicated that both EC_50_ for PD-1 receptor occupancy and E_max_ for IL-2 concentration were not significantly associated with PFS and OS (Fig. 3E-F).

**Fig. 3.**
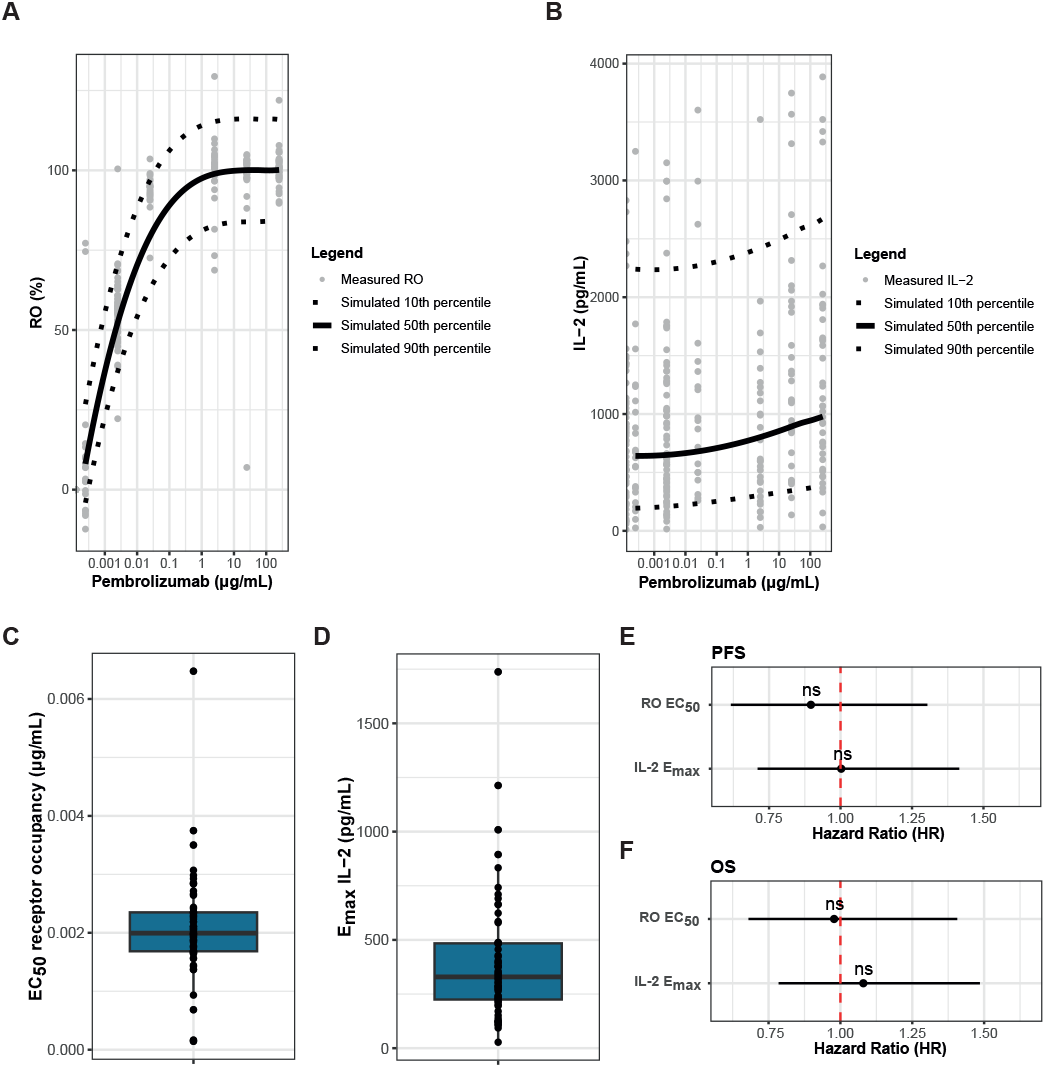
PD-1 receptor occupancy and IL-2 induction following *in vitro* stimulation are not associated with PFS and OS. Measured (**A**) PD-1 receptor occupancy (n = 60) and (**B**) IL-2 concentration (n = 61) in response to increasing pembrolizumab concentrations following *in vitro* stimulation, with corresponding *in silico* modelled concentration-response curves. Simulated (**C**) EC_50_ of PD-1 receptor occupancy (n = 60) and (**D**) E_max_ of IL-2 concentration (n = 61). Forest plots for the association of the EC_50_ of PD-1 receptor occupancy and the E_max_ of IL-2 concentration with (**E**) PFS and (**F**) OS. PD-1 = programmed cell death protein 1, IL-2 = interleukin-2, PFS = progression free survival, OS = overall survival, RO = receptor occupancy, EC_50_ = half maximal effective concentration, E_max_ = maximum effect, HR = hazard ratio, ns = not significant.

#### 3.3.2. Treatment with pembrolizumab in NSCLC patients modulates specific kinases and phenotype markers that are associated with response

To evaluate the effect of pembrolizumab therapy on the peripheral blood T lymphocyte compartment, we studied T cell signalling and phenotype at BL and after 3w and 6w of *in vivo* treatment, focussing on markers whose association with response were modulated by therapy. Forest plots, summarizing the survival analysis, of all evaluated covariates (percentages and ratios of cell subsets, expression levels of phenotype markers, and phosphorylation levels of signalling markers) at all time points are presented in Fig. S3. Before treatment initiation, higher expression of HLA-DR and CD28 on CD4+ terminal effector memory T cells (CD27-CD45RA+) (Temra), increased percentage of CD4-CD8-T cells and elevated platelet to lymphocyte ratio were negatively associated with survival, as reflected by increased HRs (HRs 1.37 – 1.86) (Fig. 4A). However, following 3w and 6w of clinical pembrolizumab treatment, these associations were no longer statistically significant, indicating that the markers were modulated by the *in vivo* pembrolizumab treatment.

**Fig. 4.**
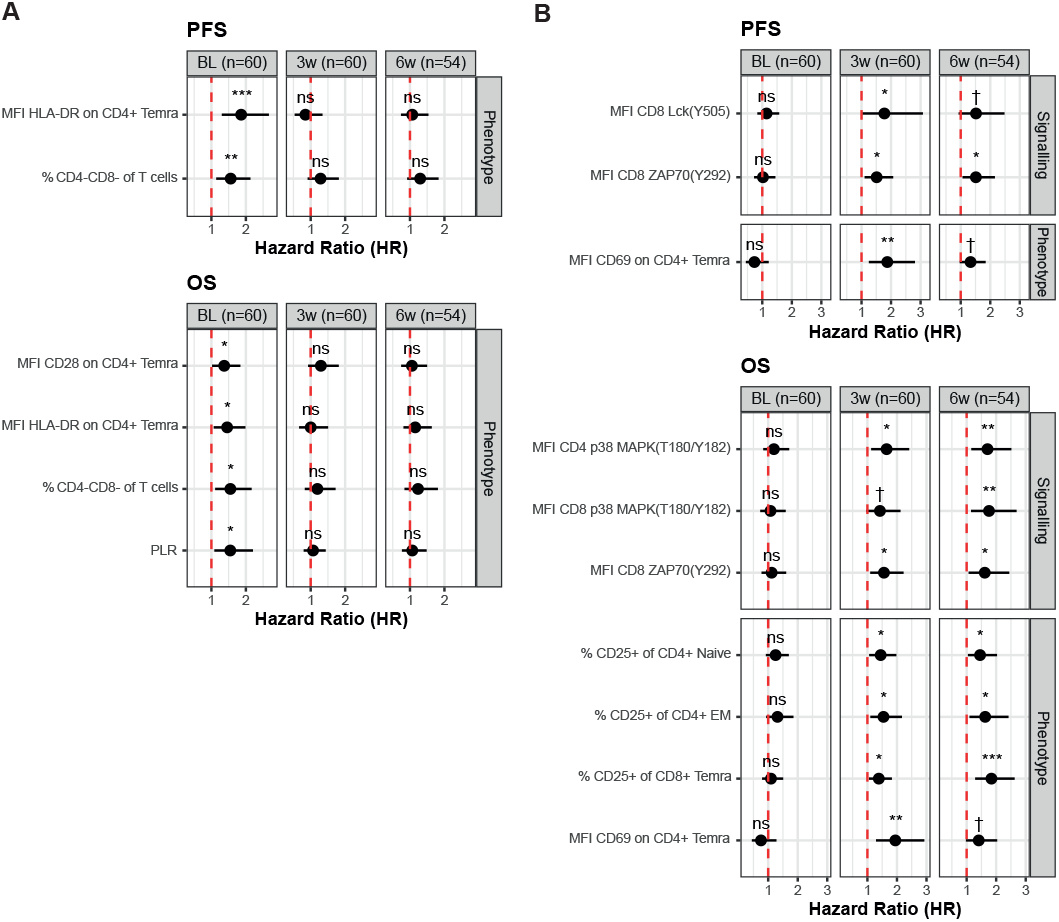
T cell signalling and phenotype markers whose association with PFS and OS are modulated by *in vivo* pembrolizumab treatment. (**A**) Hazard ratios (HRs) of markers which were significantly associated with PFS and/or OS at BL in univariate Cox regression, but lost significance after 3w and 6w of *in vivo* pembrolizumab treatment. (**B**) HRs of markers which were not significantly associated with PFS and/or OS at BL, but that became significant after 3w and 6w of *in vivo* pembrolizumab treatment. N = 54-60 patients. BL = baseline, 3w = three weeks, 6w = six weeks, PFS = progression free survival, OS = overall survival, HR = hazard ratio, Temra = terminal effector memory T cell (CD27-CD45RA+), HLA-DR = human leukocyte antigen – DR isotype, PLR = platelet to lymphocyte ratio, Lck = leukocyte-specific tyrosine kinase, ZAP70 = zeta-chain-associated protein kinase-70, MAPK = mitogen-activated protein kinase, Naive = naive T cell (CD27+CD45RA+), EM = effector memory T cell (CD27-CD45RA-), ns = not significant, † = *p* < 0.10, ^*^ = *p* < 0.05, ^**^ = *p* < 0.01, ^***^ = *p* < 0.001.

In contrast, after treatment initiation (3w and 6w), markers negatively associated with PFS and/or OS included elevated phosphorylation levels of proximal kinases Lck(Y505) and ZAP70(Y292) in CD8+ T cells, and distal kinase p38 MAPK(Thr180/Tyr182) in both CD4+ and CD8+ T cells, as well as increased levels of T cell phenotype markers CD69 expression on CD4+ Temra cells and the percentages of CD4+ naive T cells (CD27+CD45RA+), CD4+ effector memory (EM) T cells (CD27-CD45RA-) and CD8+ Temra cells expressing CD25 (HRs 1.33 – 1.95) (Fig. 4B). These signalling and phenotype markers reflect treatment-dependent immunological modulations that associate with clinical response.

#### 3.3.3. Pretreatment in vitro pembrolizumab-dependent T cell signalling profiles associate with OS

To investigate whether the pembrolizumab-dependent functional T cell phenotype, observed following 3w and 6w of *in vivo* pembrolizumab treatment, could already be detected prior to treatment initiation, we studied the effect of pembrolizumab on T cell signalling and phenotype in BL samples in an unstimulated condition or following *in vitro* stimulation in the absence or presence of pembrolizumab. When analysing all phenotype and signalling markers together by means of hierarchical clustering analysis, no distinct clustering based on the immune profile could be observed across the three *in vitro* conditions (Fig. S4 A-C).

Therefore, to focus on T cell functionality, we examined patterns of changes in the BL samples: ΔStim (stimulated minus unstimulated) and ΔPembro (stimulated in the absence versus presence of pembrolizumab), alongside unstimulated values (Unstim), focusing on T cell phosphorylation signatures. In these pretreatment samples, in the unstimulated condition, unsupervised hierarchical clustering of T cell phosphorylation profiles identified three distinct patient clusters with: 1) inactive phosphorylation profiles, 2) active phosphorylation profiles, and 3) overactive phosphorylation profiles (Fig. 5A). Overall signalling scores in the active phosphorylation profile group were significantly higher than in the inactive phosphorylation profile group (*p* < 0.0001) and significantly lower than in the overactive phosphorylation profile group (*p* < 0.0001) (Fig. 5B). However, these distinct T cell signalling profiles were not significantly associated with PFS or OS (Fig. 5C-D).

**Fig. 5.**
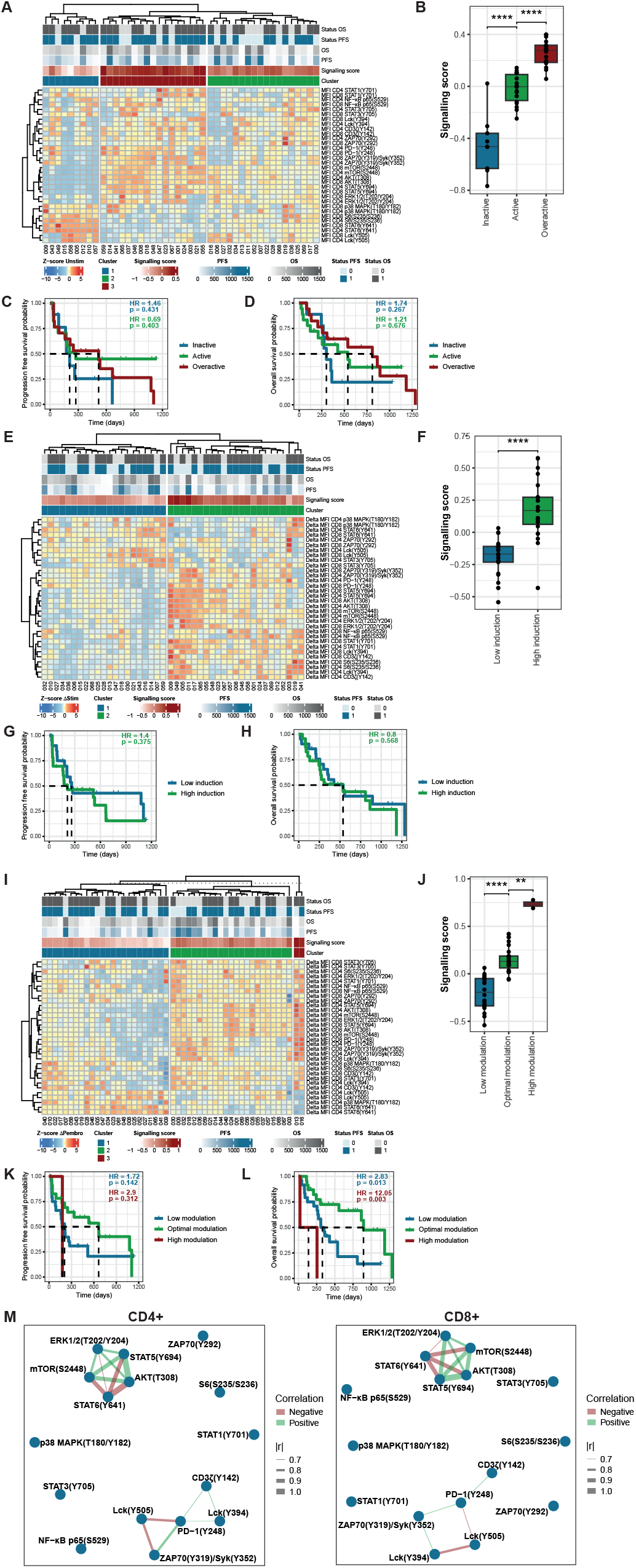
Changes in T cell phosphorylation signatures in baseline samples in response to *in vitro* pembrolizumab treatment associate with OS. (**A**) T cell signalling profiles in unstimulated samples (n = 44). (**B**) Overall signalling scores in patients with inactive, active and overactive native signalling profiles. Kaplan-Meier curves displaying (**C**) PFS and (**D**) OS for patients with inactive, active, and overactive signalling profiles in unstimulated samples. (**E**) Changes in T cell signalling profiles in response to *in vitro* stimulation in the absence of pembrolizumab compared to unstimulated samples (ΔStim; n = 44). (**F**) Overall signalling scores in patients with low and high induction in response to *in vitro* stimulation. Kaplan-Meier curves displaying (**G**) PFS and (**H**) OS for patients with low induction of T cell signalling profile, and high induction of T cell signalling profile in response to *in vitro* stimulation in the absence of pembrolizumab. (**I**) Modulation of T cell signalling profiles in response to *in vitro* stimulation in the presence of 25 µg/mL pembrolizumab compared to stimulated samples in the absence of pembrolizumab (ΔPembro; n = 49). (**J**) Overall signalling scores in low, optimal and high modulation responders. Kaplan-Meier curves displaying (**K**) PFS and (**L**) OS for patients with low, optimal, and high modulation response. (**M**) Correlation network showing spearman correlation between changes in phosphorylation levels of individual signalling markers in response to *in vitro* pembrolizumab (ΔPembro). PFS = progression free survival, OS = overall survival, STAT = signal transducer and activator of transcription, NF-κB p65 = nuclear factor-κB p65 subunit, Lck = leukocyte-specific tyrosine kinase, ZAP70 = zeta-chain-associated protein kinase-70, PD-1 = programmed cell death protein 1, Syk = spleen tyrosine kinase, mTOR = mammalian target of rapamycin, ERK = extracellular signal-regulated kinase, MAPK = mitogen-activated protein kinase, Unstim = unstimulated, HR = hazard ratio, |r| = absolute Spearman’s rank correlation coefficient (*ρ*), ^**^ = *p* < 0.01, ^****^ = *p* < 0.0001.

Following *in vitro* stimulation of the BL samples, distinct changes (ΔStim) in phosphorylation signature were observed. Hierarchical clustering analysis revealed that the three phosphorylation profiles observed in unstimulated samples submerged into two major clusters following *in vitro* stimulation: low induction of phosphorylation, and high induction of phosphorylation (Fig. 5E). This indicates that T cell stimulation via the T cell receptor (TCR) and CD28 identified immunological responders to stimulation (high induction) and non-responders (low induction) based on the phosphorylation profile. Again, overall signalling scores in the low T cell induction group were significantly higher than in the high T cell induction group (*p* < 0.0001) (Fig. 5F). However, T cell phosphorylation profiles in response to *in vitro* stimulation remained unassociated with PFS or OS (Fig. 5G-H).

Finally, when pembrolizumab was added to the *in vitro* stimulation, again distinct changes (ΔPembro) in phosphorylation signature were observed. Hierarchical clustering analysis identified three patient clusters with distinct pembrolizumab-dependent immunological response profiles: 1) low, 2) optimal, and 3) high modulation of phosphorylation in response to pembrolizumab (Fig. 5I). Pembrolizumab-dependent changes in individual signalling markers across the three modulation response groups are shown in Fig. S5. Again, the overall signalling score was significantly higher in patients with an optimal modulation response compared to patients with a low modulation response (*p* < 0.0001) and significantly lower than in patients with a high modulation response (*p* < 0.01) (Fig. 5J). Patients with low and high modulation responses exhibited the shortest median PFS and OS, along with significantly higher HRs (2.83, *p* = 0.013; and 12.05, *p* = 0.003 respectively) for OS compared to patients with an optimal modulation response (Fig. 5K-L). Taken together, this indicates that both insufficient induction, as well as excessive induction of phosphorylation in response to pembrolizumab are unfavourable for clinical outcomes, demonstrating that a well-balanced immune response is essential for treatment success.

To explore mechanistic relationships underlying pembrolizumab-dependent changes in phosphorylation levels of the individual signalling markers, we performed correlation network analysis in both CD4+ and CD8+ T cells. These analyses revealed a module of proximal T cell signalling markers, including PD-1(Y248), Lck(Y394), CD3ζ(Y142) and ZAP70(Y319)/Syk(Y352), that were highly correlated (Fig. 5M). Likewise, a module of distal signalling markers, including AKT(T308), mTOR(S2448), ERK1/2(T202/Y204) and STAT5(Y694), exhibited strong correlation. These high correlations imply that the signalling events are tightly co-regulated, so when PD-1 inhibition alters one of these phosphorylation sites, the others tend to respond in a coordinated manner. Interestingly, Lck(Y505) was negatively correlated to the proximal module and likewise, STAT6(Y641) was negatively correlated to the distal module, suggesting antagonistic relationships in response to PD-1 blockade. Patients exhibiting low pembrolizumab-dependent modulation of phosphorylation predominantly induced an Lck(Y505)/STAT6(Y641) response, whereas patients with optimal and high pembrolizumab-dependent modulation of phosphorylation induced a response in the proximal and distal modules (Fig. 5M), indicating distinct mechanistic response patterns among patients, that may underly differential clinical outcomes.

## 4. Discussion

PD-1 inhibitors remain the cornerstone of treatment for advanced-stage NSCLC, even though a durable response is limited to only a small proportion of patients. A major challenge lies in the identification of patients likely to benefit from therapy. This study describes the functional immune phenotype of T cells in response to pembrolizumab to gain deeper insight into the mechanisms underlying treatment response and to identify early-response biomarkers. Our findings reveal that pretreatment measurement of *in vitro* therapeutic modulation of T cell phosphorylation signatures can be of added value in clinical response prediction.

(Pre)clinical studies with pembrolizumab by Dulos *et al*. [23], Patnaik *et al*. [24] and our own group [9] demonstrated a pembrolizumab concentration-dependent increase in IL-2 production. Accordingly, we initially evaluated IL-2 concentration as a surrogate marker for T cell effector function and PD-1 receptor occupancy as a measure for target engagement. *In silico* simulations demonstrated a consistent gap between the EC_50_ values for PD-1 receptor occupancy and IL-2 concentration, confirming our earlier findings. [9] Therefore, it can be argued that receptor occupancy-based dose development of novel PD-1 inhibitors, as frequently employed, might not be the optimal method. [25] Notably, we observed inter-patient variability in both the EC_50_ for PD-1 receptor occupancy, as well as the E_max_ for IL-2 concentration. However, neither parameter was significantly associated with PFS or OS, suggesting that, although IL-2 has been used as a response monitor by Dulos et al. and Patnaik et al., IL-2 induction alone is insufficient to predict treatment outcome, underscoring the need for more reliable biomarkers.

To address this, we next examined T cell signalling and phenotype in peripheral blood in patients at BL and following 3w and 6w of *in vivo* pembrolizumab treatment. Pretreatment, we found that a high platelet to lymphocyte ratio, expression of HLA-DR and CD28 on CD4+ Temra cells, and CD4-CD8-T cell frequencies were associated with poor clinical outcomes. Consistently, a high platelet to lymphocyte ratio was also indicative of shorter OS in previous studies. [26, 27] Interestingly, HLA-DR and CD28 are potentially indicating functionally responsive CD4+ Temra cells that are theoretically beneficial for an anti-tumour immune response. Likewise, Lu *et al*. showed that CD4-CD8-T cells inhibited proliferation and invasion of human pancreatic cancer cells and CD4-CD8-tumour infiltrating lymphocytes have shown anti-tumour cytotoxicity against various cancer cell lines. [28, 29] Thus, their presence in peripheral blood may imply that they are not trafficking to the tumour microenvironment, limiting their therapeutic impact. Taken together, above mentioned immunopharmacodynamic markers at BL showed associations with clinical outcomes, thus implying their potential utility in identifying treatment response prior to treatment initiation.

In contrast, after treatment initiation, markers in peripheral blood in patients that were associated with clinical outcomes, included signalling markers Lck(Y505) and ZAP70(Y292) in CD8+ T cells, and p38 MAPK(Thr180/Tyr182) in both CD4+ and CD8+ T cells, as well as phenotypic markers CD69 expression on CD4+ Temra cells and the percentages of CD4+ naive T cells, CD4+ EM T cells and CD8+ Temra cells expressing CD25. As described by Hurkmans *et al*., the kinome is a potential mirror of the anti-tumour activity of the immune system. [30] Phosphorylation of Lck(Y505) maintains Lck in an inactive conformation, thereby preventing TCR signalling, while ZAP70(Y292) phosphorylation recruits inhibitory molecules that dampen downstream signalling. [15-17] Additionally, while p38 MAPK is important for T cell activation in cytokine-activated T cells, it has also been implicated as a negative regulator in TCR-activated T cells and is associated with T cell senescence. [18-20, 31] As phosphorylation at these sites is known to inhibit T cell activation, increased phosphorylation levels in CD8+ T cells may reveal ineffective pembrolizumab treatment in these patients. Unexpectedly, this inhibitory signalling profile did not align with the phenotype markers observed on CD4+ T cell subsets. CD69 and CD25 are typically upregulated following TCR engagement [32], yet their increased expression was negatively associated with clinical outcomes. This discrepancy warrants further investigation. Besides, while CD4+ T cells contribute to anti-tumour immunity, CD8+ T cells are considered the primary targets of ICI. [33] Therefore, the observed inhibitory signalling profile in these cells following 3w/6w of *in vivo* pembrolizumab treatment is remarkable, as this may reflect a dysfunctional T cell phenotype that does not translate into effective anti-tumour immunity.

Although 3w and 6w post-treatment initiation represent early time points, biomarkers that can be detected prior to therapy initiation are of even greater clinical value. To explore this, we applied an *ex vivo* immunopharmacological approach assessing pembrolizumab immuno-pharmacodynamics. Specifically, we evaluated T cell signalling and phenotype in BL samples following *in vitro* stimulation, in the absence or presence of pembrolizumab, to determine whether the pembrolizumab-dependent T cell activation phenotype observed *in vivo*, could already be detected prior to treatment initiation.

Interestingly, cluster analysis of T cell phosphorylation profiles revealed clusters of patients with distinct phosphorylation signatures across all three *in vitro* conditions. To quantify this, we calculated an overall signalling score that effectively summarised the phosphorylation profiles. Strikingly, while static (Unstim) and stimulated (ΔStim) T cell phosphorylation signatures were not associated with clinical outcome, the addition of pembrolizumab to the *in vitro* stimulation uncovered phosphorylation signatures significantly associated with OS, with patients exhibiting an optimal modulation response demonstrating a significantly longer OS than those with low or high modulation responses. Taken together, these findings demonstrate that both insufficient and excessive pembrolizumab-induced phosphorylation of T cell signalling markers are detrimental for treatment response. Hence, the pembrolizumab-dependent phosphorylation profile may serve as a promising biomarker for ICI therapy. The overall signalling score was significantly different in the optimal modulation response group, compared to the low and high response groups. Future studies should optimise this score by selecting key contributing markers and establishing clinically relevant cut-off values for the translation into a diagnostic biomarker.

The two patients exhibiting high modulation of phosphorylation in response to pembrolizumab highlight the delicate balance between effective T cell activation and overstimulation, which may impair T cell function. Such exaggerated T cell activation could reflect hyper-progression, a phenomenon marked by accelerated tumour growth following ICI therapy initiation. [34, 35] Proposed mechanisms underlying hyper-progression include excessive Th1/Th17-mediated inflammatory responses, as well as enhanced regulatory T cell (Treg) activity. Notably, Arroyo-Olarte *et al*. [36] demonstrated that effector Treg stability and function is enhanced by impaired STAT6 signalling, which was also observed in these two patients. This finding may help explain the markedly short survival observed in both patients, underscoring the importance of recognising atypical immune activation patterns that may signal adverse responses to ICI therapy.

Interestingly, correlation network analysis of pembrolizumab-dependent changes in phosphorylation levels revealed a proximal and a distal T cell signalling module with highly correlated markers, implying they form coherent signalling modules that collectively respond to PD-1 inhibition. The proximal module reflects the signalling response following T cell stimulation through the TCR, which is subject to auto-inhibition via the PD-1 pathway. We successfully profiled phosphorylation of the intracellular tail of the PD-1 receptor using the anti-PD-1(Y248) antibody developed by Bardhan *et al*. [14], who proposed that phosphorylated PD-1 in tumour biopsies could serve as a predictive biomarker for responsiveness to anti-PD-1 therapy. Contributing to the overall signalling score, PD-1(Y248) phosphorylation may also serve as a predictive biomarker in an *in vitro* stimulation setting. Upon ligand binding, PD-1 signalling leads to downregulation of the TCR phosphorylation cascade through dephosphorylation of Lck(Y394), thereby disrupting downstream phosphorylation events involving ZAP70 and CD3ζ. [37, 38] Notably, Lck(Y505), the inhibitory phosphorylation site of Lck, was negatively correlated with the proximal signalling module, consistent with its role in suppressing Lck activity. The distal signalling module reflects coordinated downstream signalling processes. [37, 39] Interestingly, STAT5 phosphorylation (positively) and STAT6 phosphorylation (negatively) were strongly correlated with this distal module, suggesting their direct involvement in the downstream signalling pathways modulated by PD-1 engagement. Distinct phosphorylation patterns of STAT proteins in response to pembrolizumab were found to discriminate between the T cell phosphorylation profiles identified in the clustering analysis, potentially reflecting underlying T helper (Th) cell subtype differentiation. [21, 40] Among optimal modulation responders, pembrolizumab-dependent modulation of STAT1 and STAT3 phosphorylation was observed, indicative of Th1 and Th17-driven immune responses. Conversely, in low responders, pembrolizumab-dependent modulation of STAT6 phosphorylation was observed, consistent with a Th2-skewed response. These findings suggest that Th1 and Th17-driven T cell responses are favourable for a durable response to ICI therapy, whereas a Th2-driven response is not favourable for treatment outcome. This observation aligns with previous work by Dulos *et al*., who demonstrated that pembrolizumab enhances Th1 and Th17 responses while suppressing Th2 responses and is consistent with existing literature, despite the reported divergent roles of Th17 cells in anti-tumour immunity. [22, 23, 41]

Our study has limitations. First, conclusions regarding the high modulation response group were based on data obtained from only two patients. Validation in a larger patient cohort will be essential to confirm these observations. Second, Th cell subtypes could only be inferred indirectly, based on STAT phosphorylation patterns. To strengthen these findings, future studies should incorporate additional markers, such as lineage-defining transcription factors, surface proteins, and cytokine profiles, to enable more accurate characterisation of Th cell differentiation.

In conclusion, our study revealed that pembrolizumab pharmacodynamics, based on phenotype and phospho-proteome, are likely associated with clinical outcomes in NSCLC patients. Importantly, our findings suggest that hyperresponsive immune activation is not beneficial; a balanced immune response is critical for treatment success. Strikingly, we demonstrated that pembrolizumab-dependent modulation of T cell phosphorylation can be linked to disease outcome and that an overall signalling score could serve as a potential predictive biomarker for therapeutic efficacy of ICIs. These insights support further development of the signalling score and prospective validation as a predictive biomarker for response to ICIs.

## Data Availability

All data produced in the present study are available upon reasonable request to the authors.

## Declarations

### Ethics approval and consent to participate

All patients participated in the prospective DEDICATION-1 trial (ClinicalTrials.gov ID: NCT04909684). This study was conducted in accordance with the Medical Research Involving Human Subjects Act (WMO) and received approval from the Medical Ethics Committee Oost-Nederland (METC Oost-Nederland, Nijmegen, The Netherlands). Written informed consent was obtained from all participants prior to inclusion.

### Consent for publication

Not applicable.

### Availability of data and materials

All data in the current study are available upon reasonable request.

### Competing interests

RTH has received grant funding, consultancy fees and payment for lectures. MVDH has received grant funding and payment for educationals/webinars. MVDH participates in the advisory boards of Abbvie, Amgen, AstraZeneca, Bayer, BMS, Boehringer Ingelheim, Janssen, Lilly, Merck, MSD, Novartis, Pfizer, Roche, Sanofi, Takeda and is chair of NVALT studies foundation and chair of section oncology NVALT. MVDH is local PI of clinical trials at AstraZeneca, BMS, GSK, Novartis, Merck, Roche, Takeda, Mirati, Abbvie, MSD, Merck, Amgen, Boehringer Ingelheim, Pfizer. All payments received by MVDH were made to the institution.

### Funding

This work was supported by Stichting Treatmeds.

### Authors’ contributions

Conceptualization and methodology: JV, RTH, BP, MVDH, HK, RS. Investigation: JV, EVR. Formal analysis: JV, RTH, HK, RS. Supervision: RTH, HK, RS. Writing–original draft: JV. Writing–review and editing: JV, RTH, BP, MVDH, HK, RS. Visualization: JV. Project administration: RTH, BP, MVDH. Funding acquisition: RTH, BP, MVDH. All authors read and approved the final manuscript.

## Acknowledgements

We thank prof. Vassiliki A. Boussiotis for providing the anti-PD-1(Y248) antibody. We further thank Bram van Cranenbroek, Laurien Waaijer, Manon Kolkman, and Annemijn Arns for their help with flow cytometry and sample processing, and Marieke Slangewal, Theodoros Basiakos and Lisanne Pranger for their help as students. The following are members of the DEDICATION consortium (in alphabetical order): Barlo N.^1^, Berk Y.^2^, Bijsmans A.R.^3^, Citgez E.^4^, Claessens N.J.M.^5^, Dumoulin D.W.^6^, Geraedts E.^7^, Hendriks L.^8^, Herder G.J.M.^9^, Hiltermann T.J.N.^10^, Jacobs W.^11^, Jansen J^12^, Kroeze M.A.^13^, Lam-Wong W.Y.^14^, van Loenhout K.^15^, van der Meer F.^16^, Mulders A.C.M.^17^, Samii S.M.^18^, Smit A.A.J.^19^, Smits-van der Graaf C.A.A.^20^, Smits-Zwinkels M.A.^21^, Staal-van den Brekel A.J.^22^, Steendam C.M.J.^23^, Steens M.M.H.^24^, Tarasevych S.P.^25^, van Vollenhoven F.H.M.^26^, de Waard W.I.Q.^27^, Wachters F.M.^28^, van Walree N.C.^29^, Youssef-El Soud M.^30^.

^1^Noordwest Ziekenhuisgroep, Alkmaar, The Netherlands.

^2^Canisius Wilhelmina Ziekenhuis, Nijmegen, The Netherlands.

^3^Maasstad Ziekenhuis, Rotterdam, The Netherlands.

^4^Medisch Spectrum Twente, Enschede, The Netherlands.

^5^Rijnstate Arnhem, Arnhem, The Netherlands.

^6^Erasmus MC, Rotterdam, The Netherlands.

^7^Groene Hart Ziekenhuis, Gouda, The Netherlands.

^8^Maastricht UMC, Maastricht, The Netherlands.

^9^Meander Medisch Centrum, Amersfoort, The Netherlands.

^10^UMC Groningen, Groningen, The Netherlands.

^11^Martini Ziekenhuis, Groningen, The Netherlands.

^12^Ikazia Ziekenhuis, Rotterdam, The Netherlands.

^13^Streekziekenhuis Koningin Beatrix, Winterswijk, The Netherlands.

^14^Elkerliek ziekenhuis, Helmond, The Netherlands.

^15^Bravis ziekenhuis, Roosendaal, The Netherlands.

^16^Diakonessenhuis, Utrecht, The Netherlands.

^17^Ziekenhuis Gelderse Vallei, Ede, The Netherlands.

^18^Deventer Ziekenhuis, Deventer, The Netherlands.

^19^Onze Lieve Vrouw Gasthuis, Amsterdam, The Netherlands.

^20^Radboudumc, Nijmegen, The Netherlands.

^21^ZorgSaam Ziekenhuis, Terneuzen, The Netherlands.

^22^Ziekenhuisgroep Twente, Almelo, The Netherlands.

^23^Catharina Ziekenhuis, Eindhoven, The Netherlands.

^24^VieCuri Medisch Centrum, Venlo, The Netherlands.

^25^Zaans Medisch Centrum, Zaandam, The Netherlands.

^26^Frisius MC, Leeuwarden, The Netherlands.

^27^Beatrixziekenhuis, Gorinchem, The Netherlands.

^28^Gelre ziekenhuizen Zutpen, Zutphen, The Netherlands.

^29^Amphia ziekenhuis, Breda, The Netherlands.

^30^Maxima Medisch Centrum, Veldhoven, The Netherlands.

The graphical abstract and Figure 1 were created with BioRender (BioRender.com) for which the authors have a licence.

## Authors’ information

## Notes

### Clinical Trial

NCT04909684

## References

1. Herzberg B, Campo MJ, Gainor JF. Immune Checkpoint Inhibitors in Non-Small Cell Lung Cancer. Oncologist. 2017; doi:10.1634/theoncologist.2016-0189.

2. Schouten RD, Muller M, de Gooijer CJ, Baas P, van den Heuvel M. Real life experience with nivolumab for the treatment of non-small cell lung carcinoma: Data from the expanded access program and routine clinical care in a tertiary cancer centre-The Netherlands Cancer Institute. Lung Cancer. 2018; doi:10.1016/j.lungcan.2017.11.012.

3. Grizzi G, Caccese M, Gkountakos A, Carbognin L, Tortora G, Bria E, et al. Putative predictors of efficacy for immune checkpoint inhibitors in non-small-cell lung cancer: facing the complexity of the immune system. Expert Rev Mol Diagn. 2017; doi:10.1080/14737159.2017.1393333.

4. Koomen BM, Badrising SK, van den Heuvel MM, Willems SM. Comparability of PD-L1 immunohistochemistry assays for non-small-cell lung cancer: a systematic review. Histopathology. 2020; doi:10.1111/his.14040.

5. Rizvi H, Sanchez-Vega F, La K, Chatila W, Jonsson P, Halpenny D, et al. Molecular Determinants of Response to Anti-Programmed Cell Death (PD)-1 and Anti-Programmed Death-Ligand 1 (PD-L1) Blockade in Patients With Non-Small-Cell Lung Cancer Profiled With Targeted Next-Generation Sequencing. J Clin Oncol. 2018; doi:10.1200/jco.2017.75.3384.

6. Sharma P, Goswami S, Raychaudhuri D, Siddiqui BA, Singh P, Nagarajan A, et al. Immune checkpoint therapy-current perspectives and future directions. Cell. 2023; doi:10.1016/j.cell.2023.03.006.

7. Trontzas IP, Syrigos KN. Immune Biomarkers for Checkpoint Blockade in Solid Tumors: Transitioning from Tissue to Peripheral Blood Monitoring and Future Integrated Strategies. Cancers. 2025; doi: 10.3390/cancers17162639.

8. Oitabén A, Fonseca P, Villanueva MJ, García-Benito C, López-López A, Garrido-Fernández A, et al. Emerging Blood-Based Biomarkers for Predicting Immunotherapy Response in NSCLC. Cancers (Basel). 2022; doi: 10.3390/cancers14112626.

9. Verdonk JDJ, Piet B, Ter Heine R, van den Heuvel MM, Smeets RL, Koenen H. Ex vivo pembrolizumab pharmacology for personalized PD-1 inhibitor therapy reveals a critical gap between receptor occupancy and T cell functionality. Int Immunopharmacol. 2025; doi:10.1016/j.intimp.2025.114754.

10. Lala M, Li TR, de Alwis DP, Sinha V, Mayawala K, Yamamoto N, et al. A six-weekly dosing schedule for pembrolizumab in patients with cancer based on evaluation using modelling and simulation. Eur J Cancer. 2020; doi:10.1016/j.ejca.2020.02.016.

11. Thurber GM, Schmidt MM, Wittrup KD. Antibody tumor penetration: transport opposed by systemic and antigen-mediated clearance. Adv Drug Deliv Rev. 2008; doi:10.1016/j.addr.2008.04.012.

12. Waaijer LA, van Cranenbroek B, Koenen H. OMIP-112: 42-Parameter (40-Color) Spectral Flow Cytometry Panel for Comprehensive Immunophenotyping of Human Peripheral Blood Leukocytes. Cytometry A. 2025; doi:10.1002/cyto.a.24927.

13. Shchelokov D, Demin O, Jr. Receptor occupancy assessment and interpretation in terms of quantitative systems pharmacology: nivolumab case study. MAbs. 2023; doi:10.1080/19420862.2022.2156317.

14. Bardhan K, Aksoylar HI, Le Bourgeois T, Strauss L, Weaver JD, Delcuze B, et al. Phosphorylation of PD-1-Y248 is a marker of PD-1-mediated inhibitory function in human T cells. Sci Rep. 2019; doi:10.1038/s41598-019-53463-0.

15. Chapman NM, Connolly SF, Reinl EL, Houtman JC. Focal adhesion kinase negatively regulates Lck function downstream of the T cell antigen receptor. J Immunol. 2013; doi:10.4049/jimmunol.1301587.

16. Salmond RJ, Filby A, Qureshi I, Caserta S, Zamoyska R. T-cell receptor proximal signaling via the Src-family kinases, Lck and Fyn, influences T-cell activation, differentiation, and tolerance. Immunol Rev. 2009; doi:10.1111/j.1600-065X.2008.00745.x.

17. Wang H, Kadlecek TA, Au-Yeung BB, Goodfellow HE, Hsu LY, Freedman TS, et al. ZAP-70: an essential kinase in T-cell signaling. Cold Spring Harb Perspect Biol. 2010; doi:10.1101/cshperspect.a002279.

18. González-Osuna L, Fukada SY, Hernández-Cáceres MP, Luz-Crawford P, Cortez C, Rojas C, et al. p38 mitogen-activated protein kinase drives senescence in CD4(+) T lymphocytes and increases their pathological potential. Immun Ageing. 2025; doi:10.1186/s12979-025-00526-8.

19. Kumar S, Principe DR, Singh SK, Viswakarma N, Sondarva G, Rana B, et al. Mitogen-Activated Protein Kinase Inhibitors and T-Cell-Dependent Immunotherapy in Cancer. Pharmaceuticals (Basel). 2020; doi:10.3390/ph13010009.

20. Li C, Beavis P, Palfreeman AC, Amjadi P, Kennedy A, Brennan FM. Activation of p38 mitogenactivated protein kinase is critical step for acquisition of effector function in cytokine-activated T cells, but acts as a negative regulator in T cells activated through the T-cell receptor. Immunology. 2011; doi:10.1111/j.1365-2567.2010.03345.x.

21. Owen KL, Brockwell NK, Parker BS. JAK-STAT Signaling: A Double-Edged Sword of Immune Regulation and Cancer Progression. Cancers (Basel). 2019; doi:10.3390/cancers11122002.

22. Basu A, Ramamoorthi G, Albert G, Gallen C, Beyer A, Snyder C, et al. Differentiation and Regulation of T(H) Cells: A Balancing Act for Cancer Immunotherapy. Front Immunol. 2021; doi:10.3389/fimmu.2021.669474.

23. Dulos J, Carven GJ, van Boxtel SJ, Evers S, Driessen-Engels LJ, Hobo W, et al. PD-1 blockade augments Th1 and Th17 and suppresses Th2 responses in peripheral blood from patients with prostate and advanced melanoma cancer. J Immunother. 2012; doi:10.1097/CJI.0b013e318247a4e7.

24. Patnaik A, Kang SP, Rasco D, Papadopoulos KP, Elassaiss-Schaap J, Beeram M, et al. Phase I Study of Pembrolizumab (MK-3475; Anti-PD-1 Monoclonal Antibody) in Patients with Advanced Solid Tumors. Clin Cancer Res. 2015; doi:10.1158/1078-0432.Ccr-14-2607.

25. Felip E, Moreno V, Morgensztern D, Curigliano G, Rutkowski P, Trigo JM, et al. First-in-human, open-label, phase 1/2 study of the monoclonal antibody programmed cell death protein-1 (PD-1) inhibitor cetrelimab (JNJ-63723283) in patients with advanced cancers. Cancer Chemother Pharmacol. 2022; doi: 10.1007/s00280-022-04414-6.

26. Diem S, Schmid S, Krapf M, Flatz L, Born D, Jochum W, et al. Neutrophil-to-Lymphocyte ratio (NLR) and Platelet-to-Lymphocyte ratio (PLR) as prognostic markers in patients with non-small cell lung cancer (NSCLC) treated with nivolumab. Lung Cancer. 2017; doi:10.1016/j.lungcan.2017.07.024.

27. Zhou Y, Liu X, Wu B, Li J, Yi Z, Chen C, et al. AGR, LMR and SIRI are the optimal combinations for risk stratification in advanced patients with non-small cell lung cancer following immune checkpoint blockers. Int Immunopharmacol. 2025; doi:10.1016/j.intimp.2025.114215.

28. Lu J, Huang C, He R, Xie R, Li Y, Guo X, et al. CD4(-)/CD8(-) double-negative tumor-infiltrating lymphocytes expanded from solid tumor tissue suppress the proliferation of tumor cells in an MHC-independent way. J Cancer Res Clin Oncol. 2023; doi:10.1007/s00432-023-04823-x.

29. Lu Y, Hu P, Zhou H, Yang Z, Sun YU, Hoffman RM, et al. Double-negative T Cells Inhibit Proliferation and Invasion of Human Pancreatic Cancer Cells in Co-culture. Anticancer Res. 2019; doi:10.21873/anticanres.13795.

30. Hurkmans DP, Verdegaal EME, Hogan SA, de Wijn R, Hovestad L, van den Heuvel DMA, et al. Blood-based kinase activity profiling: a potential predictor of response to immune checkpoint inhibition in metastatic cancer. J Immunother Cancer. 2020; doi:10.1136/jitc-2020-001607.

31. Smith PL, Piadel K, Dalgleish AG. Directing T-Cell Immune Responses for Cancer Vaccination and Immunotherapy. Vaccines (Basel). 2021; doi:10.3390/vaccines9121392.

32. Poloni C, Schonhofer C, Ivison S, Levings MK, Steiner TS, Cook L. T-cell activation-induced marker assays in health and disease. Immunol Cell Biol. 2023; doi:10.1111/imcb.12636.

33. Raskov H, Orhan A, Christensen JP, Gögenur I. Cytotoxic CD8(+) T cells in cancer and cancer immunotherapy. Br J Cancer. 2021; doi:10.1038/s41416-020-01048-4.

34. Denis M, Duruisseaux M, Brevet M, Dumontet C. How Can Immune Checkpoint Inhibitors Cause Hyperprogression in Solid Tumors? Front Immunol. 2020; doi:10.3389/fimmu.2020.00492.

35. Zhao LP, Hu JH, Hu D, Wang HJ, Huang CG, Luo RH, et al. Hyperprogression, a challenge of PD-1/PD-L1 inhibitors treatments: potential mechanisms and coping strategies. Biomed Pharmacother. 2022; doi:10.1016/j.biopha.2022.112949.

36. Arroyo-Olarte RD, Rivera-Rugeles A, Nava-Lira E, Sánchez-Barrera Á, Ledesma-Soto Y, Saavedra R, et al. STAT6 controls the stability and suppressive function of regulatory T cells. Eur J Immunol. 2023; doi:10.1002/eji.202250128.

37. Retnakumar SV, Chauvin C, Bayry J. The implication of anti-PD-1 therapy in cancer patients for the vaccination against viral and other infectious diseases. Pharmacol Ther. 2023; doi:10.1016/j.pharmthera.2023.108399.

38. Wu CS, Liu FC, Lin SC, Chyuan IT. Regulation of T cell receptor (TCR) signaling by tyrosine phosphatases: Recent advances and implication for therapeutic approach in autoimmune diseases. J Formos Med Assoc. 2025; doi:10.1016/j.jfma.2025.04.023.

39. Hu X, Li J, Fu M, Zhao X, Wang W. The JAK/STAT signaling pathway: from bench to clinic. Signal Transduct Target Ther. 2021; doi:10.1038/s41392-021-00791-1.

40. Tay RE, Richardson EK, Toh HC. Revisiting the role of CD4(+) T cells in cancer immunotherapynew insights into old paradigms. Cancer Gene Ther. 2021; doi:10.1038/s41417-020-0183-x.

41. Lee J, Lozano-Ruiz B, Yang FM, Fan DD, Shen L, González-Navajas JM. The Multifaceted Role of Th1, Th9, and Th17 Cells in Immune Checkpoint Inhibition Therapy. Front Immunol. 2021; doi:10.3389/fimmu.2021.625667.

