## Supplementary materials for "A pretreatment T cell signalling score identifies clinical pembrolizumab response in non-small cell lung cancer patients"

**Table S1. List of monoclonal antibodies used for spectral flow cytometry.**

|  | Specificity | Fluorochrome | Clone | Producer | Cat. nr. |
| --- | --- | --- | --- | --- | --- |
| **Surface panel** | CD3 | Spark Blue 550 | SK7 | BioLegend | 344852 |
|  | CD4 | Brilliant Blue 515 | SK3 | BD | 566912 |
|  | CD8 | PerCP | SK1 | BioLegend | 344708 |
|  | CD14 | Spark Violet 538 | S18004B | BioLegend | 399213 |
|  | CD19 | PE-Fire 640 | HIB19 | BioLegend | 302274 |
|  | CD25 | Brilliant Violet 605 | M-A251 | BioLegend | 356142 |
|  | CD27 | APC-Fire 810 | QA17A18 | BioLegend | 393214 |
|  | CD28 | BUV496 | CD28.2 | BD | 741168 |
|  | CD38 | Brilliant Violet 650 | HB-7 | BioLegend | 356620 |
|  | CD45RA | PE-Fire 700 | HI100 | BioLegend | 304172 |
|  | CD69 | RB780 | FN50 | BD | 568754 |
|  | CD127 | Brilliant Violet 570 | A019D5 | BioLegend | 351308 |
|  | CD274 | APC | MIH1 | Thermo Fisher | 17-5983-42 |
|  | CD279 | Brilliant Violet 785 | EH12.2H7 | BioLegend | 329930 |
|  | HLA-DR | Pacific Blue | Immu-357 | Beckman Coulter | B36291 |
|  | IgG4 Fc | PE | HP6025 | SouthernBiotech | 9200-09 |
|  | FVD | Zombie NIR | - | Biolegend | 423105 |
| **Phosflow panel 1** | CD3 | Spark NIR 685 | SK7 | Biolegend | 344862 |
|  | CD4 | Alexa Fluor 647 | SK3 | Biolegend | 344636 |
|  | CD8 | Alexa Fluor 700 | SK1 | Biolegend | 344724 |
|  | CD14 | Spark Violet 538 | S18004B | Biolegend | 399213 |
|  | CD16 | APC-Cy7 | 3G8 | BD | 557758 |
|  | CD19 | BUV496 | SJ25C1 | BD | 612938 |
|  | CD27 | APC-Fire 810 | QA17A18 | Biolegend | 393214 |
|  | CD45RA | Brilliant Violet 510 | 5H9 | BD | 740186 |
|  | CD56 | Brilliant Violet 750 | 5.1H11 | Biolegend | 362556 |
|  | AKT(T308) | Alexa Fluor 555 | D25E6 | Cell Signaling | 77440S |
|  | ERK1/2(T202/Y204) | PE-Cy5 | 6B8B69 | Biolegend | 369514 |
|  | mTOR(S2448) | PerCP-eFluor 710 | MRRBY | ThermoFisher | 46-9718-42 |
|  | NF-κB p65(S529) | PE-CF594 | K10-895.12.50 | BD | 565447 |
|  | p38 MAPK(T180/Y182) | PE | 4NIT4KK | Invitrogen | 12-9078-42 |
|  | S6(S235/S236) | V450 | N7-548 | BD | 561457 |
|  | STAT1(Y701) | Brilliant Violet 421 | 4a | BD | 562985 |
|  | STAT3(Y705) | Alexa Fluor 488 | 4/P-STAT3 | BD | 557814 |
|  | STAT5(Y694) | PE-Cy7 | 47/Stat5(pY694) | BD | 560117 |
|  | STAT6(Y641) | PerCP-Cy5.5 | 18/P-Stat6 | BD | 561195 |
|  | FVD | Zombie NIR | - | Biolegend | 423105 |
| **Phosflow panel 2** | CD3 | Spark NIR 685 | SK7 | Biolegend | 344862 |
|  | CD4 | Alexa Fluor 647 | SK3 | Biolegend | 344636 |
|  | CD8 | Alexa Fluor 700 | SK1 | Biolegend | 344724 |
|  | CD19 | BUV496 | SJ25C1 | BD | 612938 |
|  | CD27 | APC-Fire 810 | QA17A18 | Biolegend | 393214 |
|  | CD45RA | Brilliant Violet 510 | 5H9 | BD | 740186 |
|  | CD279 | Brilliant Violet 785 | EH12.2H7 | Biolegend | 329930 |
|  | CD3ζ(Y142) | eFluor 450 | 3ZBR4S | Invitrogen | 48-2478-42 |
|  | Lck(Y394) | PE | A18002D | Biolegend | 933104 |
|  | Lck(Y505) | Alexa Fluor 488 | 4/LCK-Y505 | BD | 557879 |
|  | ZAP70(Y292) | PerCP-Cy5.5 | A16038B | Biolegend | 693810 |
|  | ZAP70(Y319)/Syk(Y352) | PE-Cy7 | 1503310 | Biolegend | 683707 |
|  | PD-1(Y248)* |  |  |  |  |
|  | FVD | Zombie NIR |  | Biolegend | 423105 |

* PD-1(Y248) was a kind gift of Beth Israel Deaconess Medical Center, Harvard Medical School, Boston [17] and was conjugated to DyLight 594 using DyLight 594 Antibody Labeling Kit (Thermo Fisher Scientific, cat. nr. 46413)

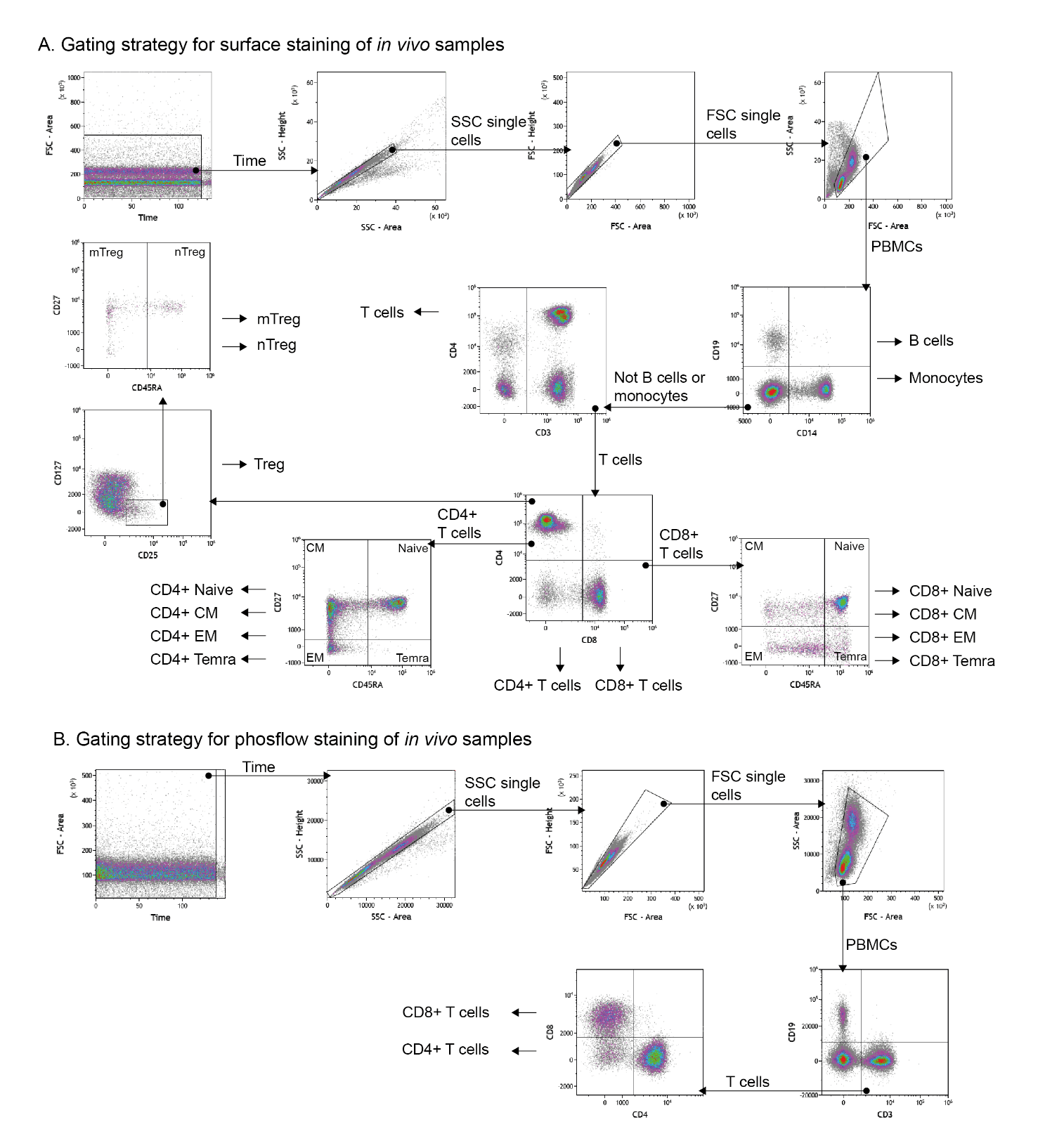
**Fig. S1** **Gating strategy**. (**A**) Surface staining and (**B**) phosflow staining of *in vivo* samples. (**C**) Surface staining and (**D**) phosflow staining of *in vitro* stimulated samples. SSC = side scatter, FSC = forward scatter, PBMCs = peripheral blood mononuclear cells, Treg = regulatory T cells, mTreg = memory Treg, nTreg = naive Treg, CM = central memory, EM = effector memory, Temra = terminal EM.

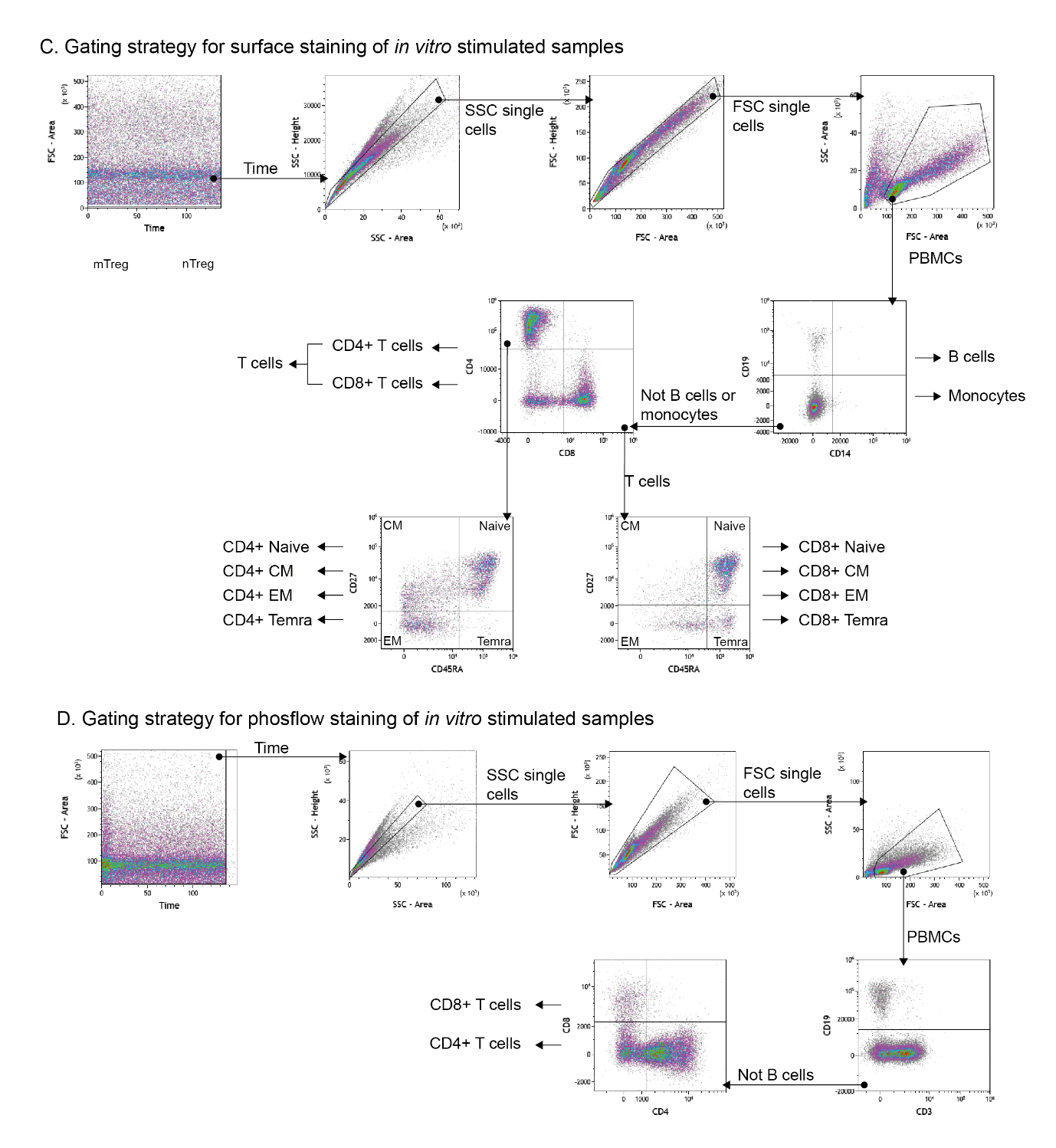

**Fig. S1** (continued).

**Table S2. Parameter estimates of in silico modelled concentration-response curves.**

|  | 1. **Receptor occupancy** | | 2. **IL-2 concentrations** | |
| --- | --- | --- | --- | --- |
|  | **Estimate** | **Inter-individual variability** (%) | **Estimate** | **Inter-individual variability** (%) |
| **Baseline** | - | - | 629.6 pg/mL  (RSE 12.66%) | 97.11  (RSE 9.17%) |
| **E_max_** | 100% (FIXED)  (RSE 8.7%) | - | 287.3 pg/mL  (RSE 33.51%) | 88.34  (RSE 13.23%) |
| **EC_50_** | 0.00184 µg/mL | 61.8  (RSE 10.7%) | 12.14 µg/mL  (RSE 169%) | - |
| **Hill** | 1.248  (RSE 5.5%) | - | 0.359  (RSE 39.35%) | - |
| **Additive residual error** | 3.47%  (RSE 9.9%) | - | - | - |
| **Proportional residual error** | 12.0%  (RSE 7.3%) | - | 14.83%  (RSE 5.79%) | - |

E_max_ = maximal effective concentration, EC_50_ = half maximal effective concentration, RSE = relative standard error of estimates.

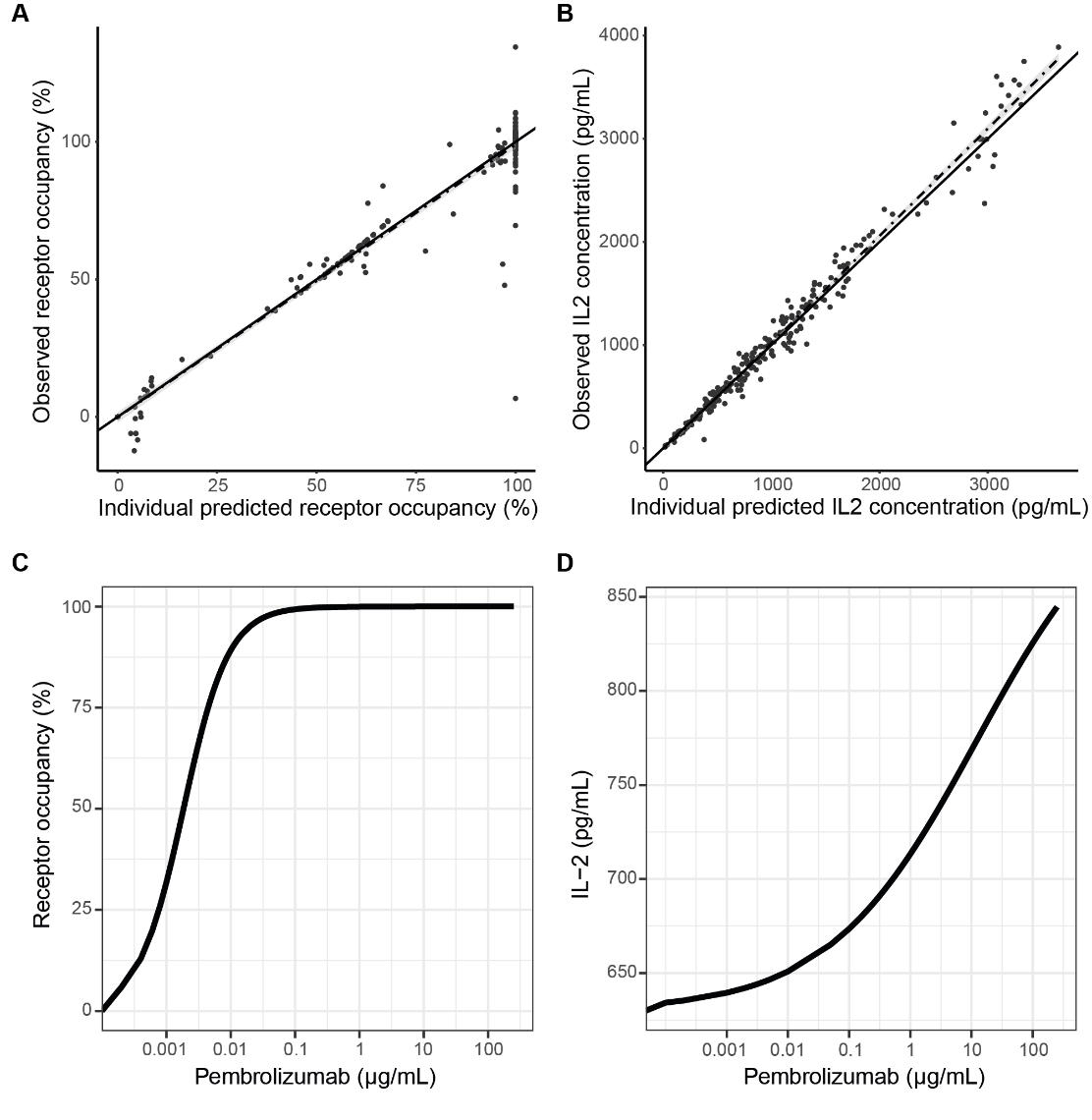

**Fig. S2 Goodness-of-fit plots and typical curves of in silico modelled receptor occupancy (A and C) and IL-2 concentrations (B and D).**

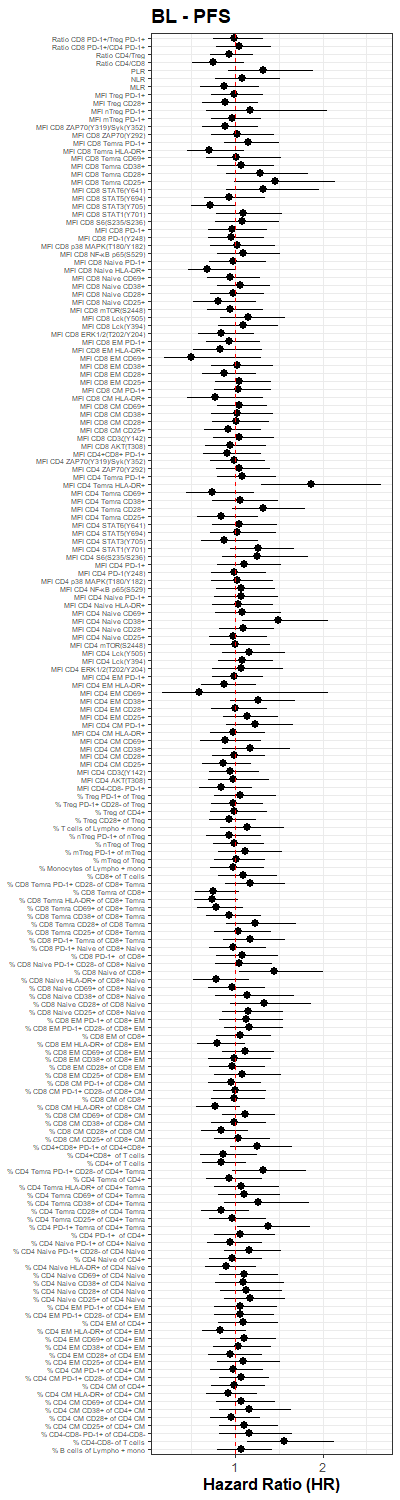

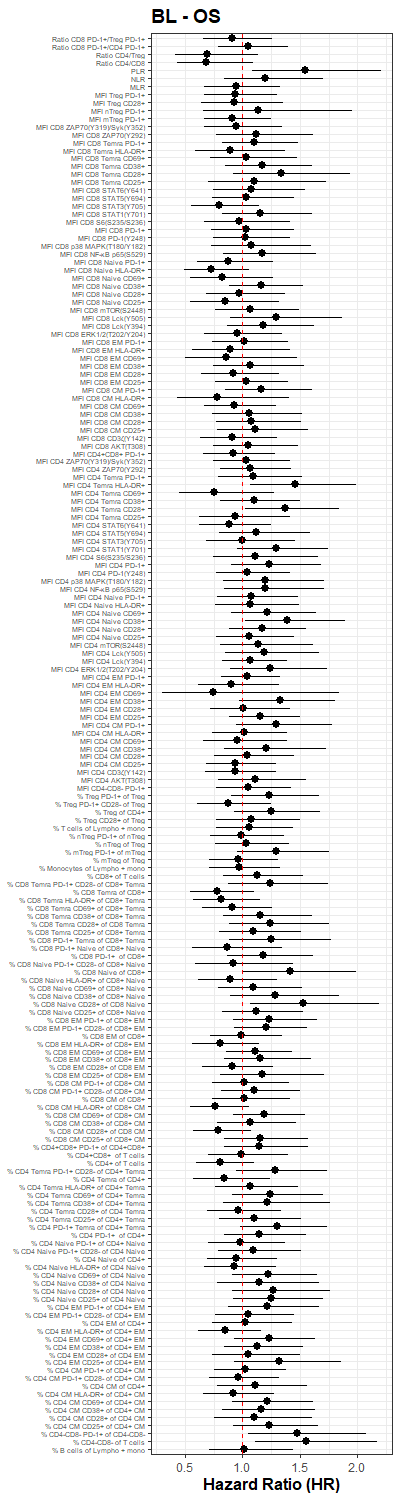

**Fig. S3 Forest plots with hazard ratios of T cell signalling and phenotype markers from BL, 3w and 6w samples.** (Caption below)

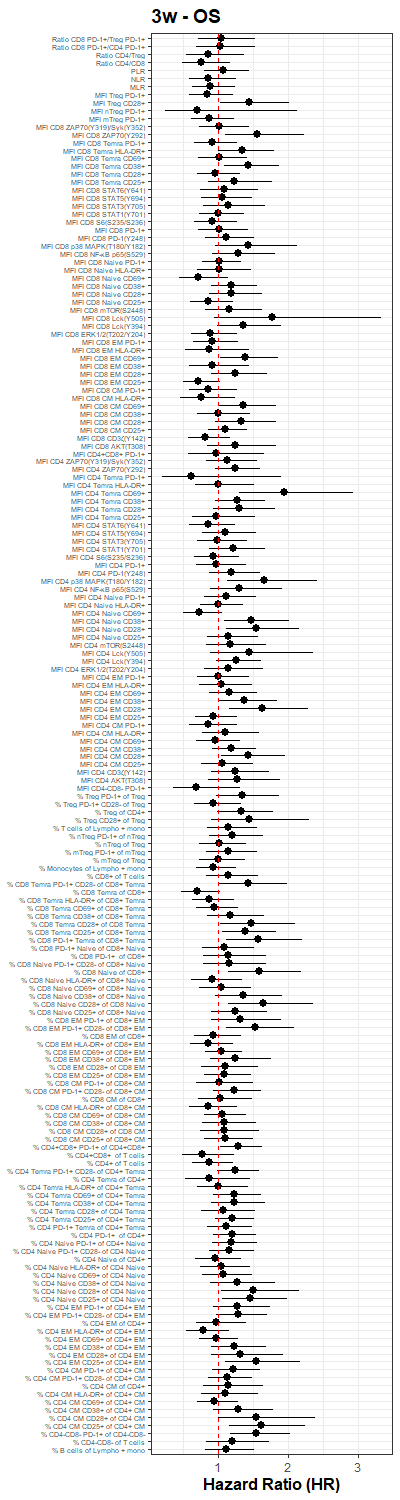

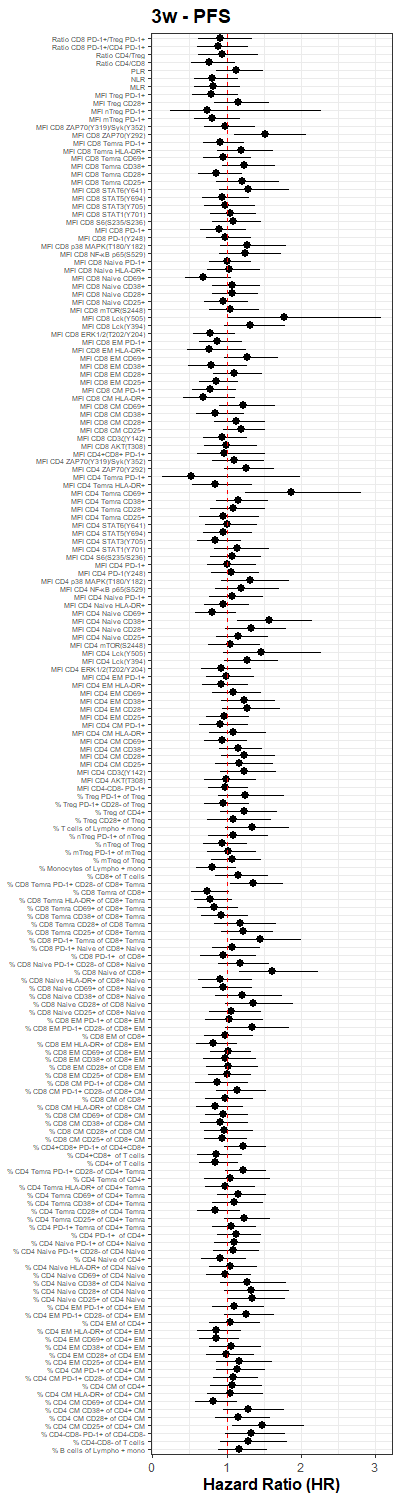
**Fig. S3** (continued).

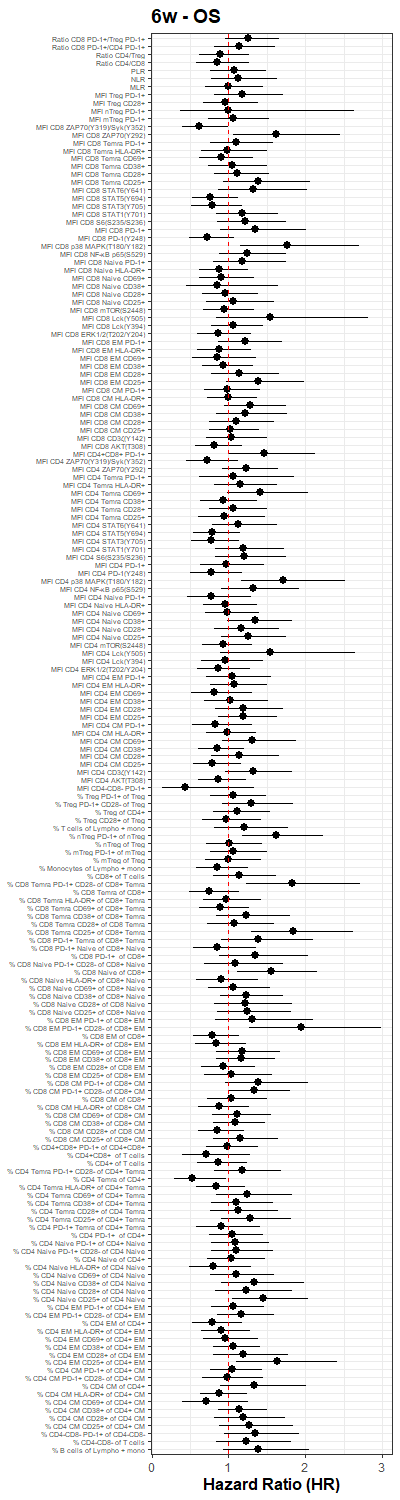

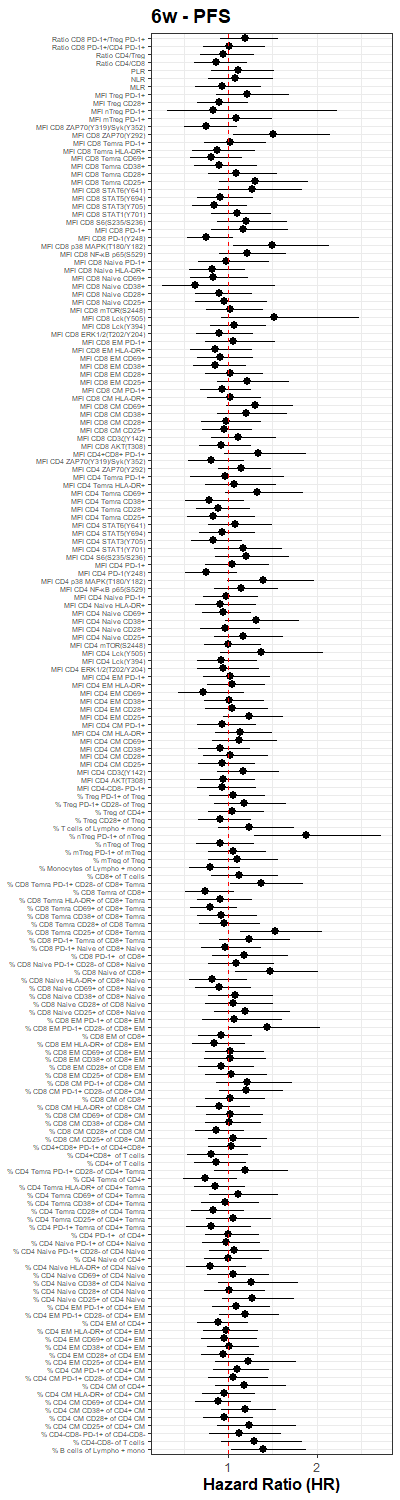
**Fig. S3** (continued).

**Fig. S3 Forest plots with hazard ratios of T cell signalling and phenotype markers from BL, 3w and 6w samples.** BL = baseline, 3w = three weeks, 6w = six weeks, PFS = progression free survival, OS = overall survival, HR = hazard ratio, MFI = mean fluorescence intensity, PD-1 = programmed cell death protein 1, Treg = regulatory T cell, PLR = platelet to lymphocyte ratio, NLR = neutrophil to lymphocyte ratio, MLR = monocyte to lymphocyte ratio, ZAP70 = zeta-chain-associated protein kinase-70, Syk = spleen tyrosine kinase, Temra = terminal effector memory T cell (CD27-CD45RA+), HLA-DR = human leukocyte antigen – DR isotype, STAT = signal transducer and activator of transcription, MAPK = mitogen-activated protein kinase, NF-κB = nuclear factor-κB p65 subunit, Naive = naive T cell (CD27+CD45RA+), mTOR = mammalian target of rapamycin, Lck = leukocyte-specific tyrosine kinase, ERK = extracellular signal-regulated kinase, EM = effector memory T cell (CD27-CD45RA-), CM = central memory T cell (CD27+CD45RA-), Lympho + mono = lymphocytes + monocytes, nTreg = naive Treg, mTreg = memory Treg.

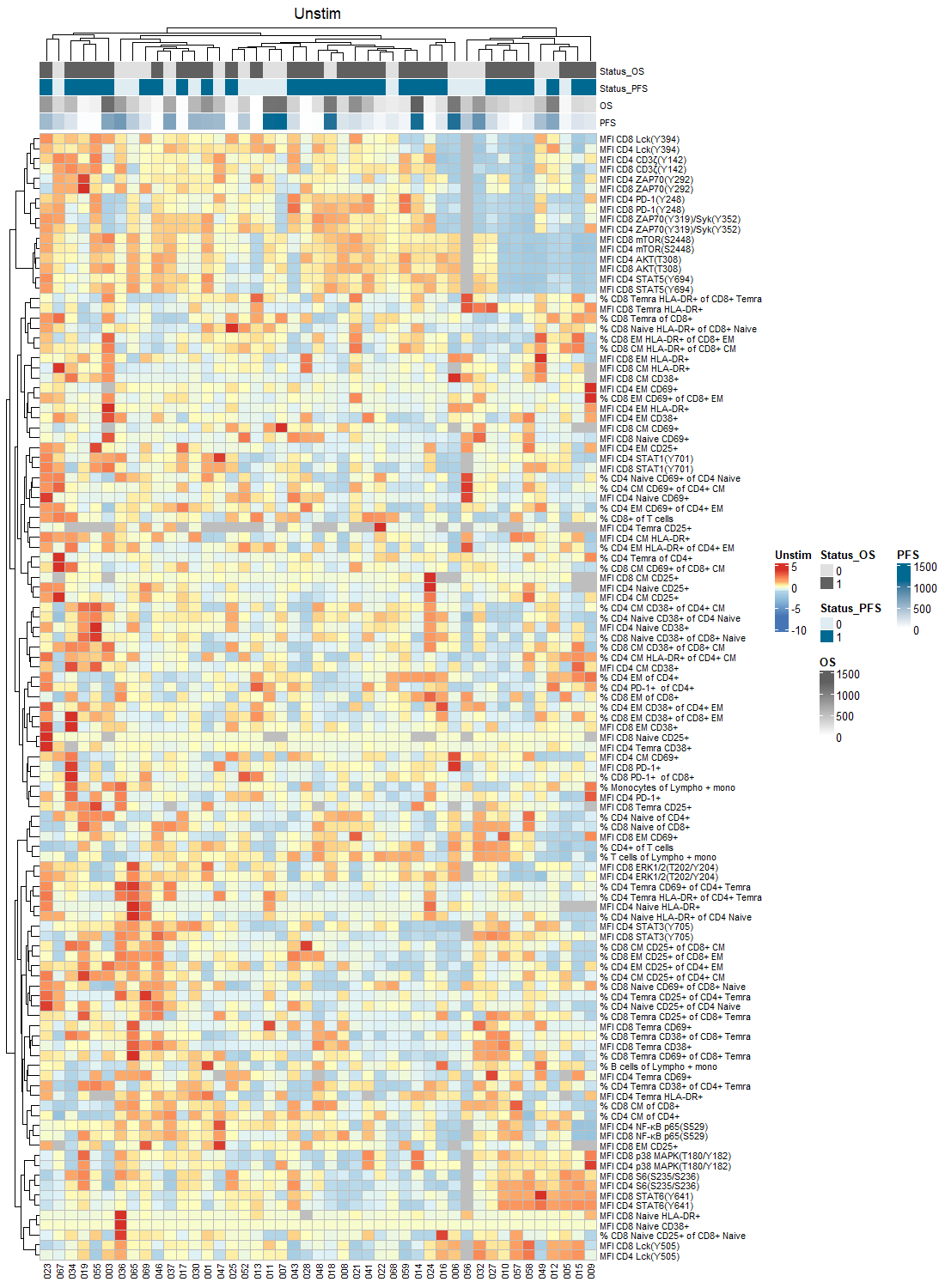

**Fig. S4 Clustering analysis of all T cell signalling and phenotype markers at BL in unstimulated samples and samples stimulated in the absence or presence of pembrolizumab.** (Caption below)

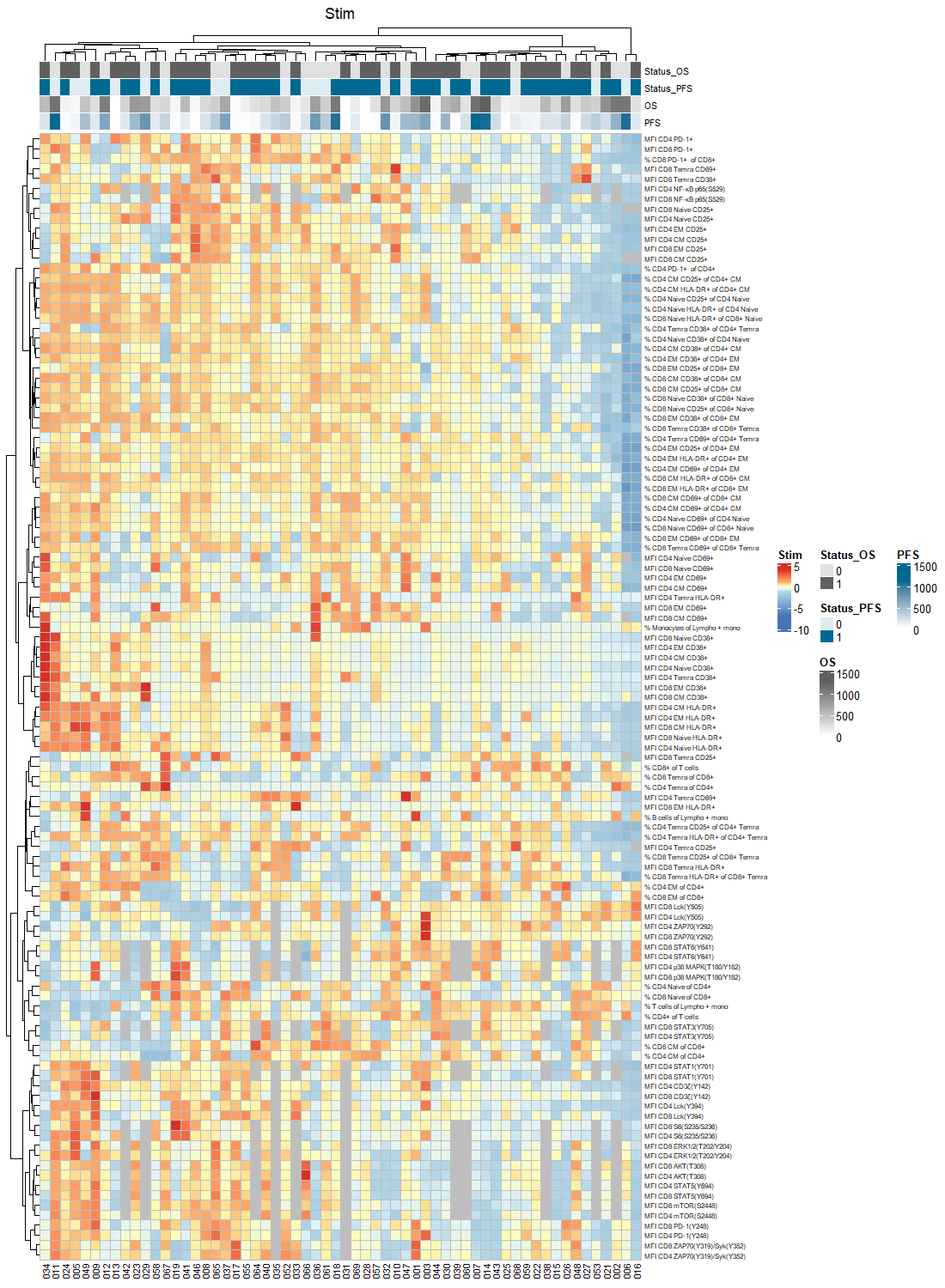

**Fig. S4** (continued).

**Fig. S4**
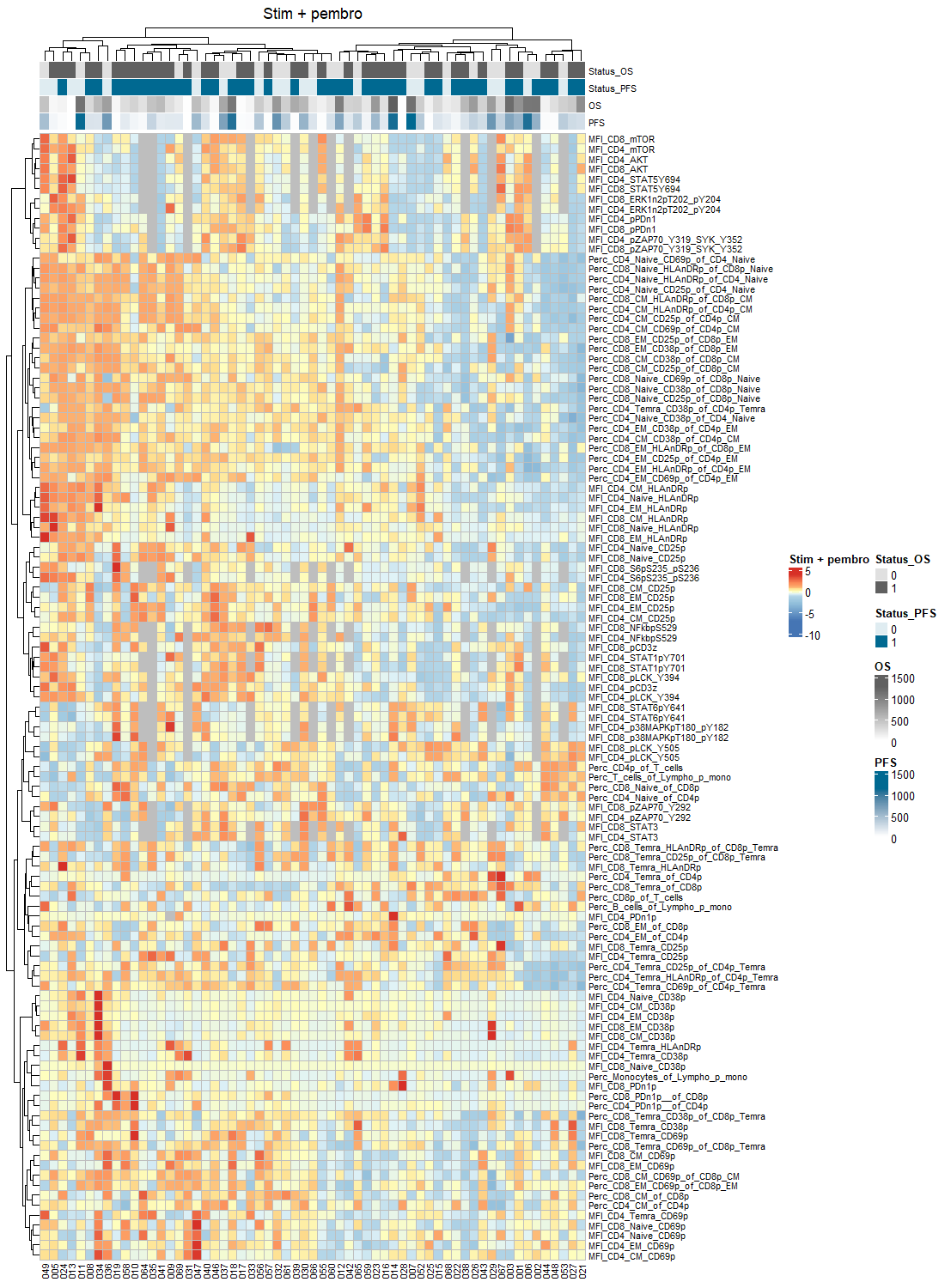
(continued).

**Fig. S4 Clustering analysis of all T cell signalling and phenotype markers at BL in unstimulated samples and samples stimulated in the absence or presence of pembrolizumab.** Phosphorylation levels (MFI) of T cell signalling markers and expression (MFI) and percentages (%) of T cell phenotype markers in unstimulated samples Unstim), samples stimulated in the absence of pembrolizumab, and samples stimulated in the presence of 2.5 or 25 ug/mL pembrolizumab (Stim + Pembro). BL = baseline, MFI = mean fluorescence intensity, OS = overall survival, PFS = progression free survival, Lck = leukocyte-specific tyrosine kinase, ZAP70 = zeta-chain-associated protein kinase-70, PD-1 = programmed cell death protein 1, Syk = spleen tyrosine kinase, mTOR = mammalian target of rapamycin, STAT = signal transducer and activator of transcription, Temra = terminal effector memory T cell (CD27-CD45RA+), HLA-DR = human leukocyte antigen – DR isotype, Naive = naive T cell (CD27+CD45RA+), EM = effector memory T cell (CD27-CD45RA-), CM = central memory T cell (CD27+CD45RA-), Lympho + mono = lymphocytes + monocytes, ERK = extracellular signal-regulated kinase, NF-κB = nuclear factor-κB p65 subunit, MAPK = mitogen-activated protein kinase.

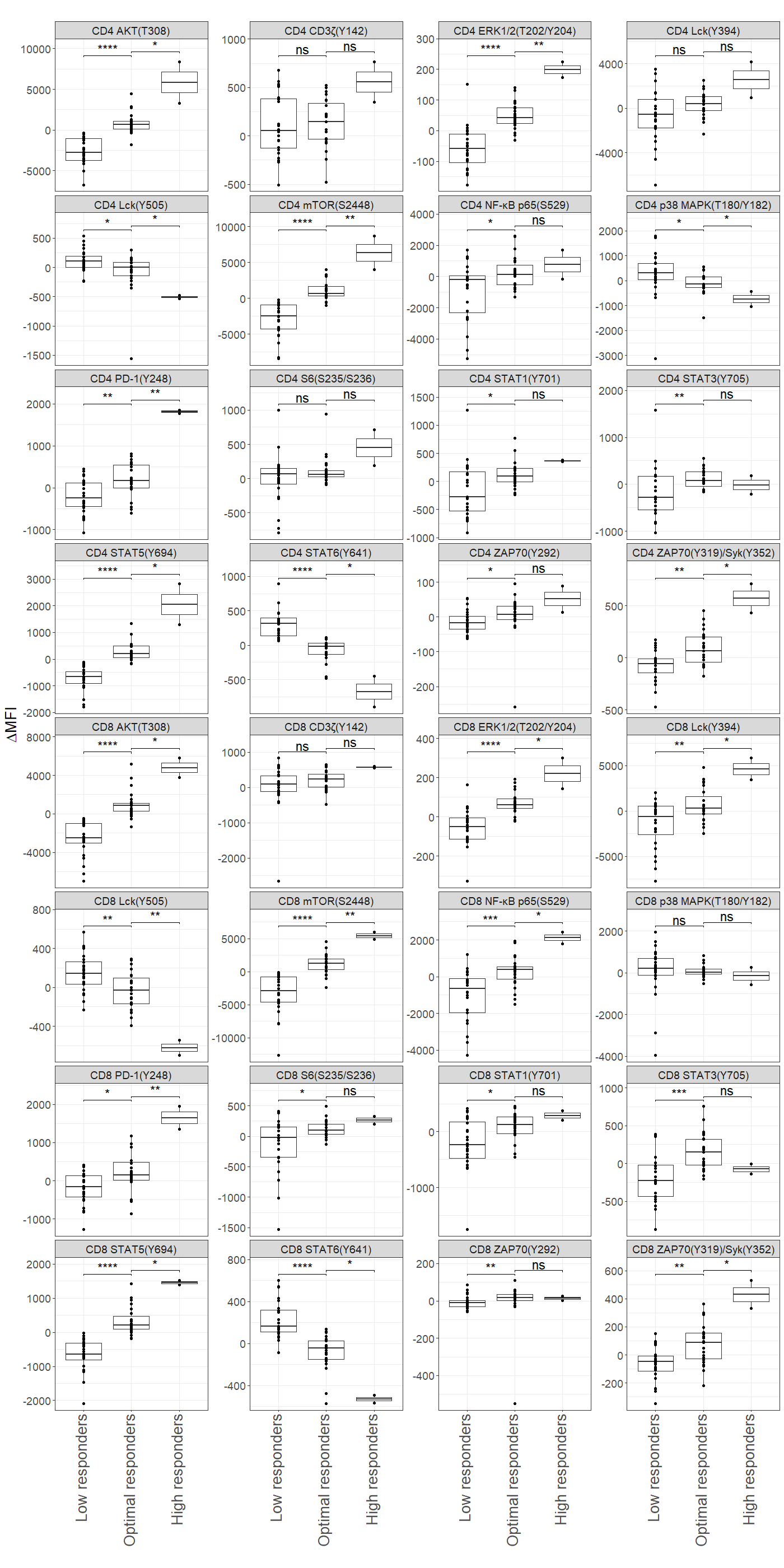
**Fig. S5 Pembrolizumab-dependent changes in phosphorylation levels of T cell signalling markers.** MFI = mean fluorescence intensity, ERK = extracellular signal-regulated kinase, Lck = leukocyte-specific tyrosine kinase, mTOR = mammalian target of rapamycin, NF-κB p65 = nuclear factor-κB p65 subunit, MAPK = mitogen-activated protein kinase, PD-1 = programmed cell death protein 1, STAT = signal transducer and activator of transcription, ZAP70 = zeta-chain-associated protein kinase-70, Syk = spleen tyrosine kinase, ns = not significant, * = p < 0.05, ** = p < 0.01, *** = p < 0.001, **** = p < 0.0001.
